# Child wasting and concurrent stunting in low- and middle-income countries

**DOI:** 10.1101/2020.06.09.20126979

**Authors:** Andrew Mertens, Jade Benjamin-Chung, John M Colford Jr, Alan E Hubbard, Mark J van der Laan, Jeremy Coyle, Oleg Sofrygin, Wilson Cai, Wendy Jilek, Sonali Rosete, Anna Nguyen, Nolan N Pokpongkiat, Stephanie Djajadi, Anmol Seth, Esther Jung, Esther O Chung, Ivana Malenica, Nima Hejazi, Haodong Li, Ryan Hafen, Vishak Subramoney, Jonas Häggström, Thea Norman, Parul Christian, Kenneth H Brown, Benjamin F. Arnold, members of the ki Child Growth Consortium

## Abstract

Sustainable Development Goal 2.2, to end malnutrition by 2030, includes elimination of child wasting, defined as weight-for-length more than 2 standard deviations below international standards. Prevailing methods to measure wasting rely on cross-sectional surveys that cannot measure onset, recovery, and persistence — key features that inform preventive interventions and disease burden estimates. We analyzed 21 longitudinal cohorts to show wasting is a highly dynamic process of onset and recovery, with incidence peaking between birth and 3 months. By age 24 months 29.2% of children had experienced at least one wasting episode, more than 5-fold higher than point prevalence (5.6%), demonstrating that wasting affects far more children than can be inferred through cross-sectional surveys. Children wasted before 6 months had faster recovery and shorter episodes than children wasted at older ages, but early wasting increased the risk of later growth faltering, including concurrent wasting and stunting (low height-for-age), increasing their risk of mortality. In diverse populations with high seasonal rainfall, population average weight-for-length varied substantially (>0.5 z in some cohorts), with the lowest mean Z-scores during the rainiest months, creating potential for seasonally targeted interventions. Our results elevate the importance of establishing interventions to prevent wasting from birth to age 6 months, likely through improved maternal nutrition, to complement current programs that focus on children ages 6-59 months.

## Introduction

Wasting is a form of undernutrition that results from a loss of muscle and fat tissue from acute malnutrition, affecting an estimated 45.4 million children under 5 years (6.7%) worldwide with over half living in South Asia.^1^ Children are considered wasted if their weight-for-length z-score (WLZ) falls 2 standard deviations below the median of international standards, and severe wasting is 3 standard deviations below the median.^2^ Wasted children have weakened immune systems,^3^ predisposing them to infections and more severe illness once infected.^4^ Wasting in very young children increases their risk of mortality (relative risk: 2.3), with mortality risk further increasing if they are also stunted and underweight, defined as length-for-age (LAZ) below –2 and weight-for-age (WAZ) below –2 (relative risk: 12.3).^5^ Longitudinal studies have shown that wasting is also associated with higher risk of stunting^6^ and poor neurocognitive development at older ages.^7, 8^ Community-based identification and management of severely wasted children (WLZ below –3), including treatment with high calorie, ready-to-use-therapeutic foods (RUTF) has proven effective,^9^ but rates of relapse and mortality remain high in this fragile subgroup.^10, 11^ Primary prevention of wasting would be preferable to treatment, but there are scant proven preventive interventions, especially in infants under 6 months. Nutritional intervention trials have had mixed success, with no evidence for reduced wasting with breastfeeding support from birth, and larger reductions from small-quantity lipid-based nutrient supplements between ages 6-24 months (14% relative reduction in wasting, 31% relative reduction in severe wasting).^9, 12, 13^

Wasting is thought to occur primarily from ages 6-24 months during the critical period when exclusive breastfeeding is no longer recommended and adequate, appropriate and safe complementary feeding is required,^14, 15^ but a more complete understanding of the epidemiology of wasting and how it varies by age is key to develop and target preventive interventions.^14, 16, 17^ Unlike the cumulative process of linear growth faltering and stunting,^18^ wasting varies considerably over time, both within-individuals and within-populations.^19, 20^ The dynamic nature of wasting means that the number of distinct episodes a child experiences may be poorly captured in cross-sectional, survey-based estimates.^21^ Furthermore, seasonal patterns of wasting onset in relation to changes in food insecurity or disease can only be measured accurately with longitudinal data.^17, 22, 23^ A synthesis of longitudinal cohort data across diverse populations provides a unique opportunity to study the timing, dynamics, and burden of wasting in infants and young children — key knowledge gaps to inform future prevention efforts.

### Pooled longitudinal analyses

Here, we report a pooled analysis of 21 longitudinal cohorts from 10 low- and middle-income countries (LMICs) in South Asia, Sub-Saharan Africa, and Latin America, which measured length and weight monthly among children 0-24 months of age. Our primary objectives were to produce the first large-scale estimates of wasting incidence and recovery and to quantify the temporal and regional variation. We also assessed the concurrence of wasting and stunting and compared estimates of wasting prevalence and incidence. We analyzed data from the Bill & Melinda Gates Foundation’s Knowledge Integration (*ki*) database, which has aggregated studies on child growth and development.^24^ Inclusion criteria were defined to select cohorts representative of general populations in LMICs with sufficiently frequent measurements to investigate the acute, dynamic nature of wasting. Cohorts were included if they: 1) conducted in low- or middle-income countries; 2) enrolled children between birth and age 24 months and measured their length and weight repeatedly over time; 3) did not restrict enrollment to acutely ill children; 4) had a median birth year of 1990 or later 5) measured anthropometry at least monthly (Extended Data Fig 1). Twenty-one cohorts met the inclusion criteria, encompassing 11,448 children and 198,154 total anthropometry measurements from birth to age 24 months (Fig 1).

**Figure 1.**
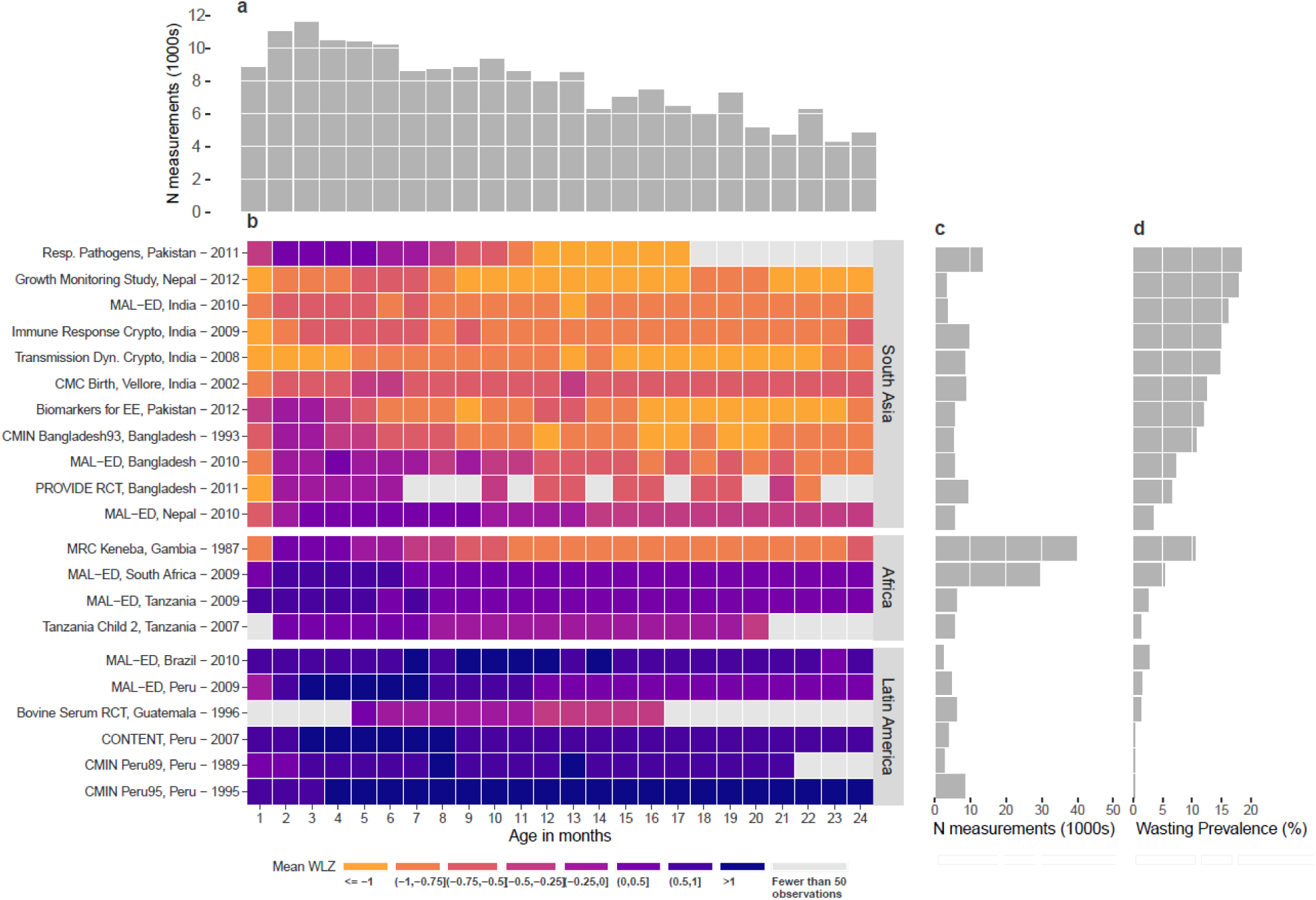
Summaries of included cohorts. (**a**) Number of observations (1000s) across cohorts by age in months. (**b**) Mean weight-for-length Z-scores (WLZ) by age in months for each included cohort. Cohorts are sorted by geographic region and overall mean weight-for-length Z-score. The country of each cohort and the start year is printed by each cohort name. (**c**) The number of observations included in each cohort. (**d**) Overall wasting prevalence by cohort, defined as proportion of measurements with WLZ < –2.

We calculated WLZ and LAZ-scores using WHO 2006 growth standards.^25^ We dropped 385 biologically implausible measurements (0.2%) of WLZ (> 5 or < –5 Z-score), 212 (0.1%) of LAZ (> 6 or < –6 Z-score), and 216 (0.1%) of WAZ (>5 or < –6), following WHO recommendations.^26^ Most included cohorts did not measure children past 24 months, so analyses focused on birth to 24 months. Cohorts ranged in size from 160 children in the TDC cohort to 2,931 children from the MRC Keneba cohort (Fig 1). Unless otherwise indicated, we conducted individual-level analyses within cohorts and then pooled cohort-specific estimates using random effects models fitted with restricted maximum likelihood estimation, a conservative approach when cohort-specific estimates are heterogeneous. Cohorts were distributed throughout South Asia, Africa, and Latin America, but did not cover entire regions (Extended Data Fig 2). Most cohorts measured children every month through age 24 months, but there was some attrition as children aged and there were four cohorts with few measurements beyond 15 months (Extended Data Fig 3). Pooled estimates at older ages could be slightly biased if unmeasured cohorts or children were systematically different from those that were included — for example, if children who were lost to follow-up were more likely to be wasted, we might have under-estimated wasting at older ages.

We assessed *ki* cohort representativeness by comparing Z-score measurements to population-based samples in Demographic and Health Surveys (DHS). Mean z-scores in *ki* cohorts were generally representative based on country-level DHS estimates, with lower WLZ (overall and by age) in Guatemala, Pakistan, and South Africa and higher WLZ in Brazil and Guatemala (Extended Data Fig 4). LAZ was generally lower in *ki* cohorts, so estimates of concurrent wasting and stunting may be higher than estimates from population-based samples, and rates of wasting recovery in early life may be higher compared with the general population.

### Age-specific patterns of wasting

Across all cohorts, mean WLZ was near –0.5 at birth and then increased over the first 6 months before decreasing until 12 months (Fig 2a). Age-specific patterns in WLZ were similar across geographic regions, but levels varied substantially by region. WLZ was markedly lower among South Asian cohorts, reflecting a much higher burden of malnutrition (Fig 2a). Children were wasted for 16,139 (8.6%) measurements, and severely wasted for 3,391 (1.8%) measurements. Wasting prevalence was highest at birth (11.9%; 95% CI: 7.0, 19.5) in contrast with prior studies that showed peak wasting prevalence between 6-24 months old (Fig 2b).^6, 27–29^ Across regions, wasting prevalence decreased from birth to three months before increasing until 12 months old; however, wasting was far more prevalent in South Asian cohorts. In South Asian cohorts – where low birthweight is common^30^– at-birth wasting prevalence was 18.9% (95% CI: 15.0, 23.7), implicating causes of poor fetal growth like maternal malnutrition, maternal morbidities, and maternal small stature as key regional drivers of wasting.^31, 32^ Severe wasting followed a similar pattern but was much rarer (Extended Data Fig 5).

**Figure 2.**
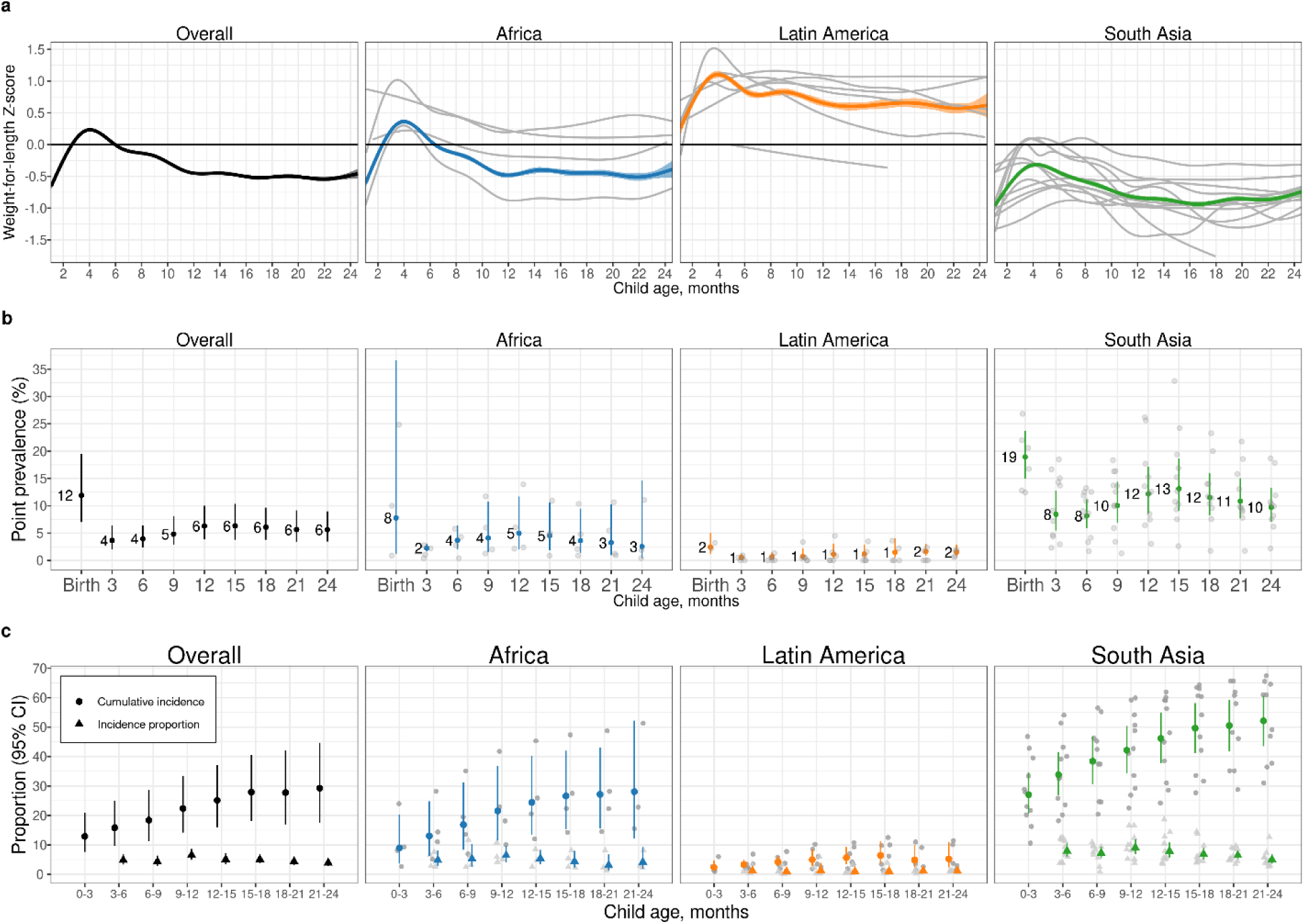
WLZ, prevalence and incidence of wasting by age and region. (**a**) Mean weight-for-length Z-score (WLZ) by age in 21 longitudinal cohorts, overall (N=21 studies; N=4,165-10,886 observations per month) and stratified by region (Africa: N=4 studies, N=1,067-5,428 observations; Latin America: N=6 studies, N= 569-1718 observations, South Asia 11 studies, N=2,382-4,286 observations). (**b**) Age-specific wasting prevalence, defined as WLZ < –2, overall (N=3,985-9,906 children) and stratified by region (Africa: 1,701-5,017 children, Latin America: N= 290-1397 children, South Asia: N=1,994-3,751 children). (**c**) Age-specific wasting incidence overall (N=6,199 -10,377 children) and stratified by region (Africa: N=2,249-5,259 children, Latin America: N=763-1,437 children, South Asia: N=3,076-3,966 children). Cumulative incidence measures the proportion of children who have ever experienced wasting since birth, while the new incident cases represent the proportion of children at risk who had an episode of wasting begin during the age period. Error bars in panels b and c are 95% confidence intervals for pooled estimates. In each panel, grey curves or points show cohort-specific estimates.

Wasting onset was highest during the first 3 months largely due to its high occurrence at birth (Fig 2c). Overall, 12.9% (95% CI: 7.6, 20.1) of all children experienced wasting by age 3 months, which accounted for almost half (47.8%) of children who ever experienced wasting in their first two years of life. Focusing on wasting prevalence alone masked how common it was for children to experience wasting in their first 24 months. After birth, up to 6.5% (95% CI: 4.9, 8.6) of children were wasted at a specific visit, but 29.2% (95% CI: 17.5, 44.7) of children experienced at least one wasting episode by 24 months of age. Cumulative incidence was highest in South Asian cohorts (52.2% [95% CI: 43.5, 60.7]), Figs 2b, c). Early onset of wasting was consistently high across countries with different levels of health spending, poverty, and under-5 mortality, with early wasting particularly high (18.3% [95% CI: 13.5, 24.4] at-birth) in the five cohorts with birth measures and a national health expenditure < 4.5% of gross domestic product (Extended Data Fig 6). WHO Child Growth Standards overestimate wasting at birth among children born preterm,^33^ though adjustment for gestational age among four cohorts with available data only reduced at-birth stunting prevalence by 0.8% and increased underweight prevalence by 0.7% (Extended Data Fig 7). Even if preterm birth were to account for some of the at-birth wasting, the small birth size, irrespective of cause, documented in this analysis raises concern because of its consequences for child growth faltering and mortality during the first 24 months.^32^

### Wasting incidence and recovery

There was high variability in WLZ across longitudinal measurements, with an average within-child standard deviation of WLZ measurements of 0.76, compared to 0.64 for LAZ and 0.52 for WAZ. We thus defined unique wasting episodes by imposing a 60-day recovery period, covering two consequent monthly measurements, where a child’s WLZ measurements needed to remain above –2 to be considered recovered and at risk for a future episode (Fig 3a). Children who were born wasted were considered to have an incident episode of wasting at birth. More than half of all first wasting episodes occurred before age 9 months (median age of first episode onset: 264 days) with later onset for recurrent episodes among children who experienced them (median age of second episode onset: 449 days, median age of third episode onset: 551 days). The mean number of wasting episodes experienced and wasting incidence at all ages was higher in South Asia, with highest incidence in the first 3 months with- or without-including episodes at birth (Fig 3b). Ultimately, most children recovered from moderate or severe wasting episodes, reinforcing its status as an acute condition. Children met our definition of “recovered” in 91.5% of episodes of moderate and 82.5% of episodes of severe wasting. Loss-to-follow-up may have affected recovery estimates because recovery can only occur in children surviving the episode. We did not have data on wasting treatment, so recovery may be more common in these highly monitored cohorts than in the general population due to treatment referrals. Across all cohorts, children recovered from 39.0% (95% CI: 34.0, 44.3) of wasting episodes within 30 days, 64.7% (95% CI: 59.3, 69.4) within 60 days, and 71.1% (95% CI: 66.1, 75.6) within 90 days, with lower recovery in South Asian cohorts compared with other regions (Fig 3c).

**Figure 3.**
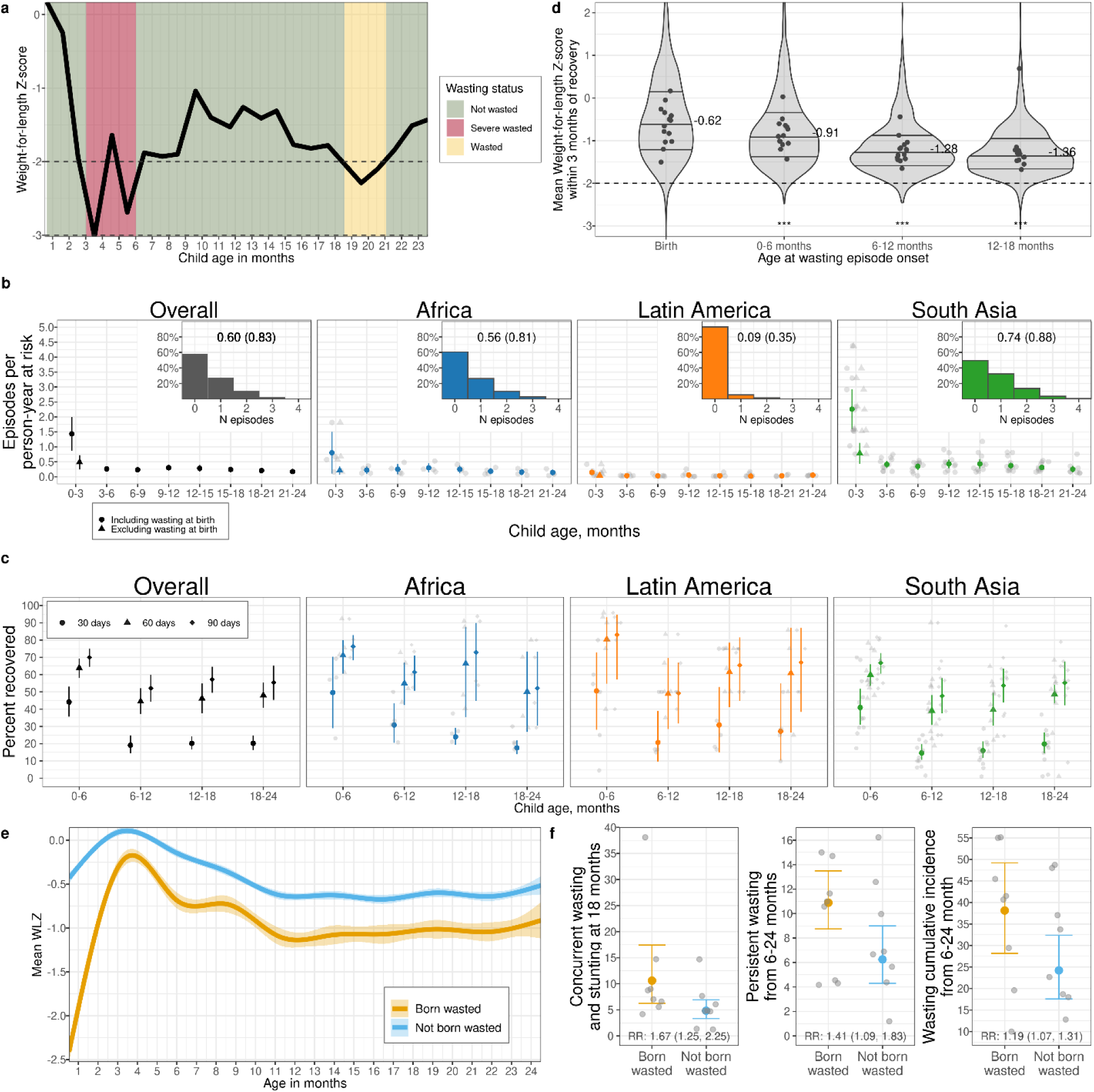
Wasting incidence rate and recovery. **(a)** Example child WLZ trajectory and wasting episode classifications. The age of wasting onset was assumed to occur halfway between a measurement of WLZ < –2 and the previous measurement of WLZ ≥ –2. Recovery from an episode of wasting or severe wasting occurred when a child had measurements of WLZ ≥ –2 for at least 60 days, with the age of recovery assumed to be halfway between the last measurement of WLZ < –2 and the first measurement of WLZ ≥ –2. **(a)** (b) Wasting incidence rate per 1,000 days at risk, stratified by age and region (Overall: N= 510,070-822,802 person-days per estimate, Africa: N=245,046-407,940 person-days, Latin America: N= 65,058-142,318 person-days, South Asia: N=182,694-315,057 person-days). Insets are histograms of the number of wasting episodes per child by region, with the distribution mean (SD) printed within the plot. (b) Percentage of children who recovered from wasting within 30, 60, and 90 days of episode onset (N=21 cohorts, 5,549 wasting episodes). (c)) The distribution of children’s mean WLZ in the three months following recovery from wasting, with inter-quartile range marked. The median WLZ is annotated, and gray points mark cohort-specific medians. Children wasted before age 6 months experienced larger improvements in WLZ compared with children wasted at older ages (p<0.001). The analysis uses 3, 686 observations of 2,301 children who recovered from wasting episodes, with 1,264 observations at birth, 628 observations from 0-6 months, 824 observations from 6-12 months, and 970 observations from 12-18 months. (a) (e) Mean WLZ by age, stratified by wasting status at birth (which includes the first measure of a child within 7 days of birth), shows that children born wasted (N= 814 children, 14,351 observations) did not catch up to children not born wasted (N= 3,355 children, 62,568 observations). (b) (f) Increased risk of multiple types of wasting after age 6 months among children born wasted (N=814), with the relative risks (RR) for born wasted versus not and 95% confidence intervals printed at the bottom (N=3,355). In panels b, d, and f, vertical lines mark 95% confidence intervals for pooled means of study specific estimates (light points).

We examined the distribution of WLZ in the three-month period after a child was considered recovered from wasting, stratified by the age at which the episode occurred. We found that children recovered to a higher WLZ following at-birth wasting or wasting episodes in the 0-6 month age window compared to episodes that occurred at older ages (Fig 3d). Regression to the mean (RTM) may explain some wasting recovery, especially the rapid WLZ gain among children born wasted (Figure 3e). However, there was catch-up growth beyond the calculated RTM effect (Extended Data Fig 8)^34^ and we required a 60-day recovery period to better capture true recovery. Additionally, placental insufficiency causes a large proportion of intrauterine growth restriction leading to birth wasting, and rapid WLZ gain after may reflect true recovery to a child’s growth potential after this constraint is removed.^35, 36^ Consistent with larger WLZ increases during recovery from wasting among younger children, a larger proportion of children recovered within 30, 60, and 90 days if the wasting episode occurred before 6 months of age (Fig 3d). Younger children had more variable WLZ (SD: 0.81 <6 months, 0.54 from 6-24 months) and shorter wasting episodes (Extended Data Fig 9). Wasting episodes were also longer at all ages in South Asian compared to African or Latin American cohorts (Extended Data Fig 9). Longer episodes among older children are consistent with higher prevalence and lower incidence compared with children <6 months.

On average, the WLZ of children born wasted did not catch up to the WLZ of children not born wasted (Fig 3e). Notably, children who were born wasted but recovered had a higher cumulative incidence of wasting after 6 months of age (38.1% cumulative incidence in children born wasted [95% CI: 28.2, 49.2] versus 24.2% cumulative incidence in children not born wasted [95% CI: 17.6, 32.4], Fig 3f). Longitudinal analyses showed a negative, reinforcing relationship between wasting and stunting: at nearly every age, wasting increased the risk of future stunting and stunting increased the risk of future wasting, with the strength of relationship increasing as children aged. This observation is consistent with longer duration of wasting episodes (Extended Data Fig 10).

### Seasonality of wasting

We joined monthly rainfall totals to cohorts by location and year., For each cohort we estimated a seasonality index based on rainfall (details in Methods). We examined both the average WLZ over the calendar year and seasonal changes with respect to rainfall, under the hypothesis that seasonal changes in food availability and infection associated with rainfall could cause seasonal wasting.^37^ Mean WLZ varied dramatically by calendar date in almost all cohorts, with a consistent minimum mean WLZ coinciding with peak rainfall (Fig 4a). When pooled across cohorts, mean WLZ was -0.16 (95% CI: -0.19, -0.14) lower during the three-month period of peak rainfall, compared to the mean WLZ in the opposite three-month period of the year, (Fig. 4b). Mean seasonal decline in WLZ was -0.27 (95% CI: -0.31, -0.24) in cohorts with a seasonality index > 0.9, but some cohorts showed seasonal WLZ declines of more than –0.5 (Fig. 4b).

**Figure 4.**
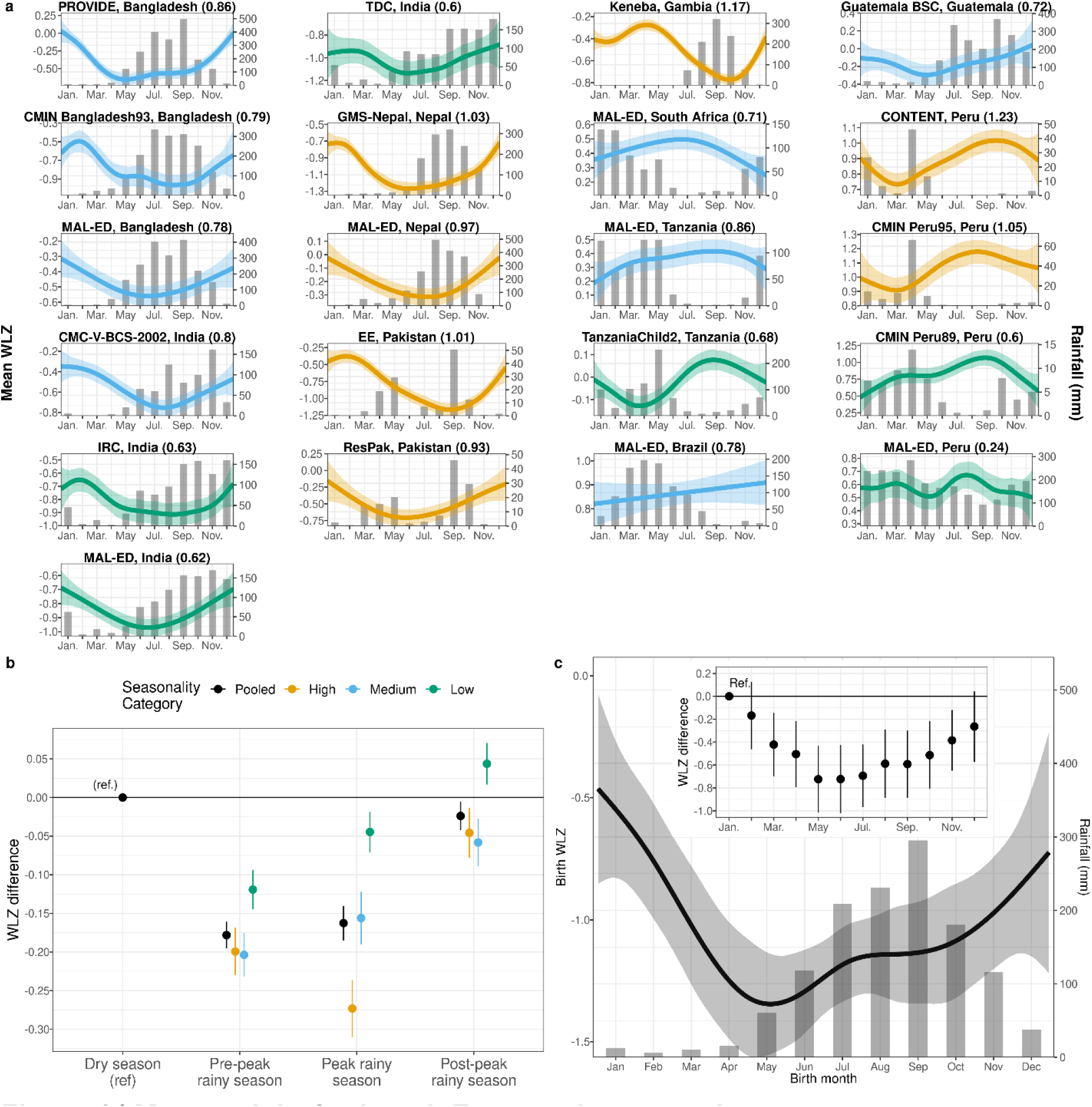
Mean weight-for-length Z-scores by age and season. **(a)** Mean WLZ by day of the year, superimposed over histograms of monthly mean rainfall over study periods, with the seasonality index printed in parentheses beside the cohort name. Means (lines) estimated with cubic splines and shaded regions indicate pointwise 95% confidence intervals. Panels are sorted by country and mean WLZ colored by high (≥ 0.9), medium (< 0.9 and ≥ 0.7), and low seasonality index categories (< 0.7). Sample sizes range from 160 children (2,545 measurements) in TDC to 2,545 (40,115 measurements) in the Keneba cohort. **(b)** Mean differences in child WLZ between quarters of the year defined around the adjacent 3-month periods with the highest mean rainfall, pooled across cohorts in panel **a**, overall (N= 21 cohorts, 2,545-40,115 observations) and by seasonality index (high: N=7 cohorts, 3,164-40,115 observations, medium: N=8 cohorts, 2,545-9,202 observations, low: N=6 cohorts, 2,741-29,518 observations). Points represent mean differences compared to the dry season and error bars represent 95% confidence intervals. **(c)** Mean WLZ at birth by month of birth estimated with a cubic spline among 1,821 children with WLZ measured at birth in 10 South Asian cohorts. The mean (line) was estimated with cubic splines and the shaded region is its pointwise 95% confidence interval. The inset plot summarizes mean differences in birth WLZ by month of birth (points) and error bars represent 95% confidence intervals.

Children who experienced seasonal wasting during the rainy season experienced longer episodes and were at higher risk of future wasting episodes. Rainy season wasting episodes lasted 7 days longer than dry season episodes (median 42 days [95% CI: 36.5 to 45.5] versus 35 days [95% CI: 33.5 to 38.5]). Incident wasting during the rainy season increased a child’s risk of experiencing another wasting episode in the following dry season (relative risk: 2.1, 95% CI: 1.3 to 3.4), and children who were wasted during the rainy season during their first year of life were 1.9 times (95% CI: 1.3 to 2.8) more likely to be wasted during the next rainy season.

South Asian cohorts had temporally synchronous rainfall, so we estimated WLZ at birth by calendar month, pooled across the 11 South Asian cohorts. Mean WLZ at birth varied by up to 0.72 z (95% CI: 0.43 to 1.02) depending on the month the child was born (range: -0.5 z to -1.3 z; Fig 4c), which suggests that seasonally-influenced maternal nutrition^38^ – likely mediated through intrauterine growth restriction or preterm birth^39^ – was a major determinant of WLZ at birth. Birth month also influenced the effect of season on WLZ trajectories that persisted through a child’s second year of life (Extended Data Fig 11), and children born in the three-month periods before or during maximum rainfall were more likely to be wasted after 6 months of age than children born in the 6 months after maximum rainfall (cumulative incidence ratio: 1.4 [95% CI: 1.1 to 1.9]).

### Persistent and concurrent growth faltering

We examined more severe forms of growth faltering, including persistent wasting and concurrent wasting and stunting, because these conditions are associated with higher mortality risk.^5^ We first identified a subset of children who experienced persistent wasting during their first 24 months. We used a pragmatic definition^40^ that classified children as persistently wasted if ≥ 50% of their WLZ measurements from birth to 24 months fell below –2. This allowed us to capture both frequent short wasting episodes or less frequent, longer episodes. Among 10,374 children with at least four measurements, 3.4% (95% CI: 2.0, 5.6) were persistently wasted. Persistent wasting over the first 24 months of life was highest in South Asia (7.2%, 95% CI: 5.1 to 10.4, Extended Data Fig 5a). Among all children wasted at birth, 10.9% were persistently wasted after 6 months (95% CI: 8.7 to 13.5) whereas 6.3% of children not born wasted who were persistently wasted after 6 months (95% CI: 4.3 to 9.0).

Finally, we examined the cumulative incidence of concurrent wasting and stunting and the timing of their overlap. Overall, 10.6% of children experienced concurrent wasting and stunting before 2 years of age (Extended data figure 5d), and a further 1.5% experienced concurrent severe wasting and stunting — far higher than point prevalence estimates in the present study (4.1% at 24 months, 95% CI: 2.6 to 6.4; Fig 5a) or the 4.3% prevalence estimated in children under 5 in LMIC from cross-sectional surveys.^41, 42^ Concurrent wasting and stunting was most common in South Asia, with the highest prevalence at age 21 months (Fig 5a), driven primarily by increases in stunting prevalence as children aged.^18^ Wasted and stunted children are also underweight, as their maximum possible weight-for-age z-score is below -2.35.^43^ Almost half of children who had prevalent growth faltering met two or more of these three growth faltering conditions, with 17-22% of all children experiencing multiple conditions after age 12 months (Fig 5b). Longitudinal analyses showed that early growth faltering predisposed children to concurrent growth faltering at older ages: children who were wasted by age 6 months were 1.8 (95% CI: 1.6 to 2.1) times and children stunted before 6 months were 2.9 (95% CI: 2.5 to 3.4) times more likely to experience concurrent wasting and stunting between ages 18-24 months. A companion article reports an in-depth investigation of key risk factors for persistent wasting and concurrent wasting and stunting, along with their consequences for future severe growth faltering and mortality.^32^

**Figure 5.**
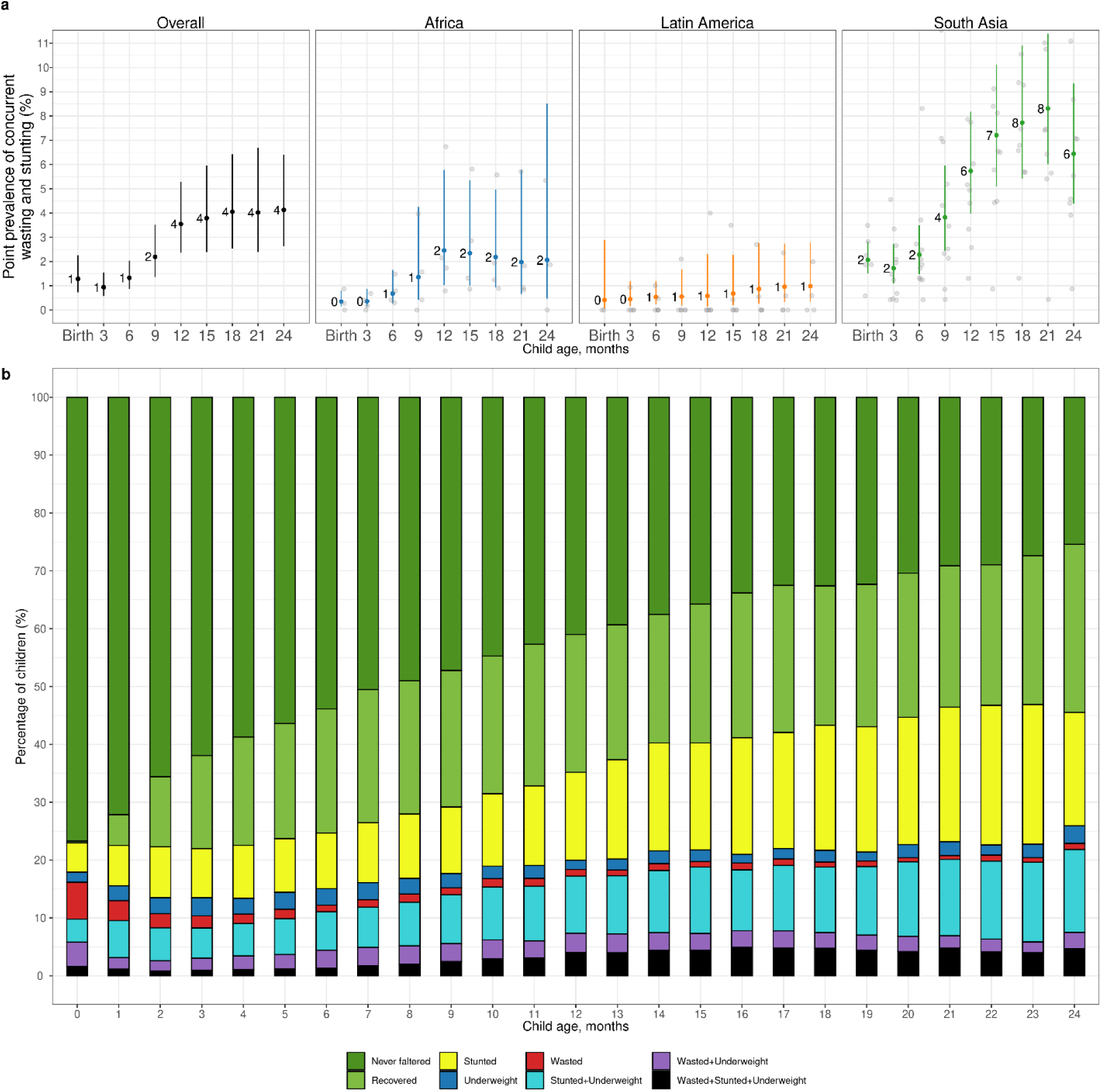
Co-occurrence of wasting, stunting, and underweight. **(a)** Age-specific prevalence of concurrent wasting and stunting overall (N=3,984 to 9,899 children) and stratified by region (Africa: N=1,799-5,014, Latin America: N=290-1,397 children, South Asia: N=1,994-3,747 children). Vertical lines mark 95% confidence intervals for pooled means across study-specific estimates (gray points). **(b)** Percentage of children classified by different measures of growth faltering, alone or combined. Children classified as “never faltered” had not previously been wasted, stunted, or underweight. Children in the “recovered” category were not wasted, stunted, or underweight but had experienced at least one of these conditions previously. All children who were wasted and stunted were also underweight. Proportions in each category were calculated within cohorts, pooled using random effects, and scaled so percentages added to 100%. The number of children contributing to each age ranges from 3,920 to 9,077 children, with 11,409 total children.

## Discussion

Nearly all large-scale studies of child wasting (including DHS) report point prevalence of wasting, which has enabled broad comparisons between populations but failed to capture the number of episodes of wasting during the course of childhood.^44, 45^ By combining information across several longitudinal cohorts, this study demonstrates that children experience far higher cumulative incidence of wasting between birth and 24 months (29.2%) than previously known. Children in all included cohorts experienced higher incidence compared with prevalence due to the episodic nature of acute malnutrition, and the pattern was most stark in South Asian cohorts where, from birth to 24 months and pooled across cohorts, 52.2% of children experienced at least one wasting episode, 7.3% were persistently wasted, and 10.1% had experienced concurrent wasting and stunting. This evidence shows that the cumulative, child-level burden of wasting is higher than cross-sectional prevalence measures would suggest, and that assessments of wasting and stunting in isolation fail to account for their joint burden, which can be substantial. This is of particular relevance in South Asia where the largest number of stunted and wasted children live, and while stunting prevalence is decreasing, wasting prevalence is _not._1,46

Our results show consistent patterns in the timing of wasting onset by age and by season, with important implications for therapeutic and preventive interventions. Wasting incidence was highest from birth to 3 months, and among children who experienced any wasting during their first 24 months, 48% experienced their first episode by 3 months. Children born wasted were far more likely to experience more growth faltering at older ages, including more severe forms like persistent wasting from 6-24 months and concurrent wasting and stunting at 18 months (Fig 3f). Age-specific patterns in prevalent wasting and concurrent wasting and stunting summarized in this analysis accord with previous reports.^6, 47–50^ To our knowledge, highest wasting incidence from birth to 3 months has not been previously described across diverse cohorts. We found higher rates of recovery among children who were wasted before 6 months, but early wasting episodes predisposed children to subsequent episodes at older ages, which tended to be longer and more likely to co-occur with stunting. These new findings suggest that the prenatal period through 6 months should be a focus for preventive wasting interventions because preventing episodes during this early window would reduce the risk of later, more severe forms of growth faltering and lower a child’s risk of all-cause mortality substantially, as we detail in a companion paper.^32^

Large seasonal changes in at-birth WLZ demonstrate a plasticity in population-level wasting whereby improvements are possible over a period of months rather than years or generations. WLZ varied dramatically by season in most cohorts, and in South Asia WLZ varied by up to 0.7 z at birth with consequences that persisted throughout the first 24 months (Fig 4c, Extended Data Fig 11). Seasonal effects plausibly extend to older ages: children born during the rainy season in rural Gambia – which coincide with the hunger season – experienced higher rates of all-cause mortality over their entire lifespan compared with children born during other times of the year. This suggests that in-utero programming of the immune system could be a significant consequence of pre-natal malnutrition.^51^ This study has not elucidated the mechanism that links seasonal rainfall with wasting, but the consistency of effect across diverse cultural and environmental contexts suggests shared mechanisms.^52^

Consistent timing with highly seasonal rainfall helps plan therapeutic programs, but seasonally targeted preventive interventions may benefit from a finer understanding of the mechanisms. In-depth analyses of some birth cohorts that contributed to the present synthesis suggest multiple mechanisms underlying seasonal wasting. Seasonal food insecurity coincides with peak energy demands during agricultural work periods, which in turn leads to undernutrition among pregnant and lactating mothers and their children.^17, 36, 51^ Women working in agriculture often have less time to spend with their children, which can reduce breastfeeding frequency and can result in early morning preparation of weaning foods that become contaminated in hot, humid conditions.^53, 54^ Weather-driven seasonal increases in infectious disease transmission could also play a role in seasonal wasting, but the effect would vary spatially and temporally and would not fully explain the consistent patterns observed here.

Sustainable Development Goal 2.2 calls to eliminate malnutrition by 2030, with child wasting as its primary indicator.^55^ The WHO global action plan on child wasting identified four key outcomes to achieve goal 2.2: reduce low birthweight, improve child health, improve child feeding, and improve treatment of wasting.^56^ Our results align with the WHO action plan but elevate the importance of improving at-birth child outcomes with a focus on both maternal support during pregnancy and nutritional supplementation in food-insecure populations for women of childbearing age, pregnant women, and children <24 months. At this time, evidence-based interventions that target this early window include balanced protein energy supplementation and iron and folic acid supplementation delivered to pregnant women and women of child-bearing age, and intermittent preventive sulfadoxine-pyrimethamine delivered to pregnant women in high malaria transmission regions of Africa.^57, 58^ For children, ages 6-24 months small-quantity lipid-based nutrient supplements have demonstrated reductions in moderate and severe wasting.^9^ If preventive or therapeutic interventions focus on children younger than 6 months, then they must integrate carefully with current recommendations for exclusive breastfeeding. In populations with highly seasonal rainfall, augmented, seasonally targeted supplementation should be considered.

## Data Availability

The data that support the findings of this study are available from the Bill and Melinda Gates Foundation Knowledge Integration project upon reasonable request.

https://child-growth.github.io/wasting/

https://github.com/child-growth/ki-longitudinal-growth

---

**Extended Data Figure 1.**
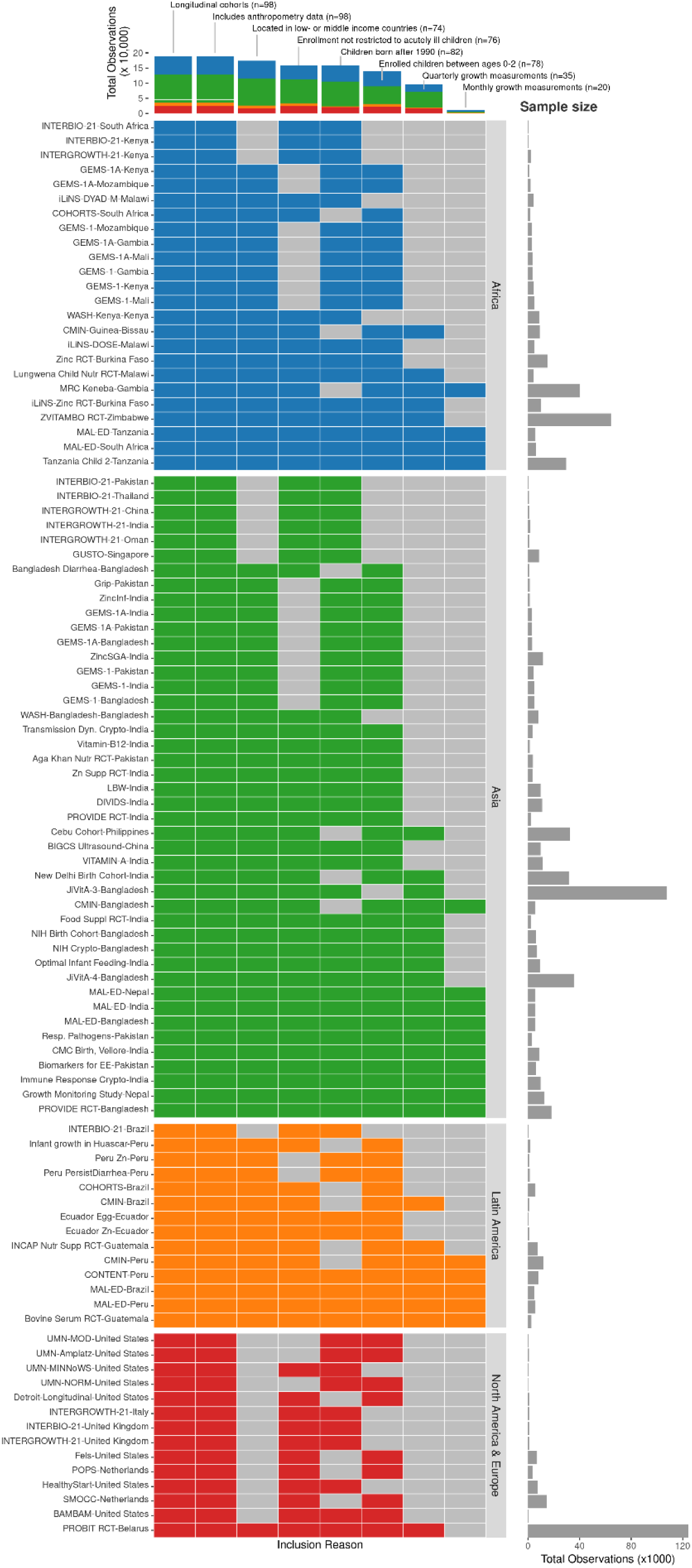
*ki* cohort selection. Analyses focused on longitudinal cohorts to enable the estimation of prospective incidence rates and growth velocity. On July 15, 2018, there were 86 longitudinal studies on GHAP. From this set, we applied five inclusion criteria to select cohorts for analysis. Our rationale for each criterion follows. (1) *Studies were conducted in lower income or middle-income countries*. Our target of inference for analyses was children in LMICs, which remains a key target population for preventive interventions. (2) *Studies measured length and weight between birth and age 24 months*. We were principally interested in growth faltering during the first two years of life including at birth, thought to be the key window for linear growth faltering ^7^. (3) *Studies did not restrict enrollment to acutely ill children*. Our focus on descriptive analyses led us to target, to the extent possible, the general population. We thus excluded some studies that exclusively enrolled acutely ill children, such as children who presented to hospital with acute diarrhea or who were severely malnourished. (4) *Studies enrolled at least 200 children*. Age-stratified incident episodes of stunting and wasting were sufficiently rare that we wanted to ensure each cohort would have enough information to estimate rates before contributing to pooled estimates. (5) *Studies collected anthropometry measurements at least every 3 months*. We limited studies to those with higher temporal resolution to ensure that we adequately captured incident episodes and recovery. We further restricted analyses of wasting incidence and recovery to cohorts with monthly measurements because of high temporal variation in WHZ within individuals.

**Extended Data Figure 2.**
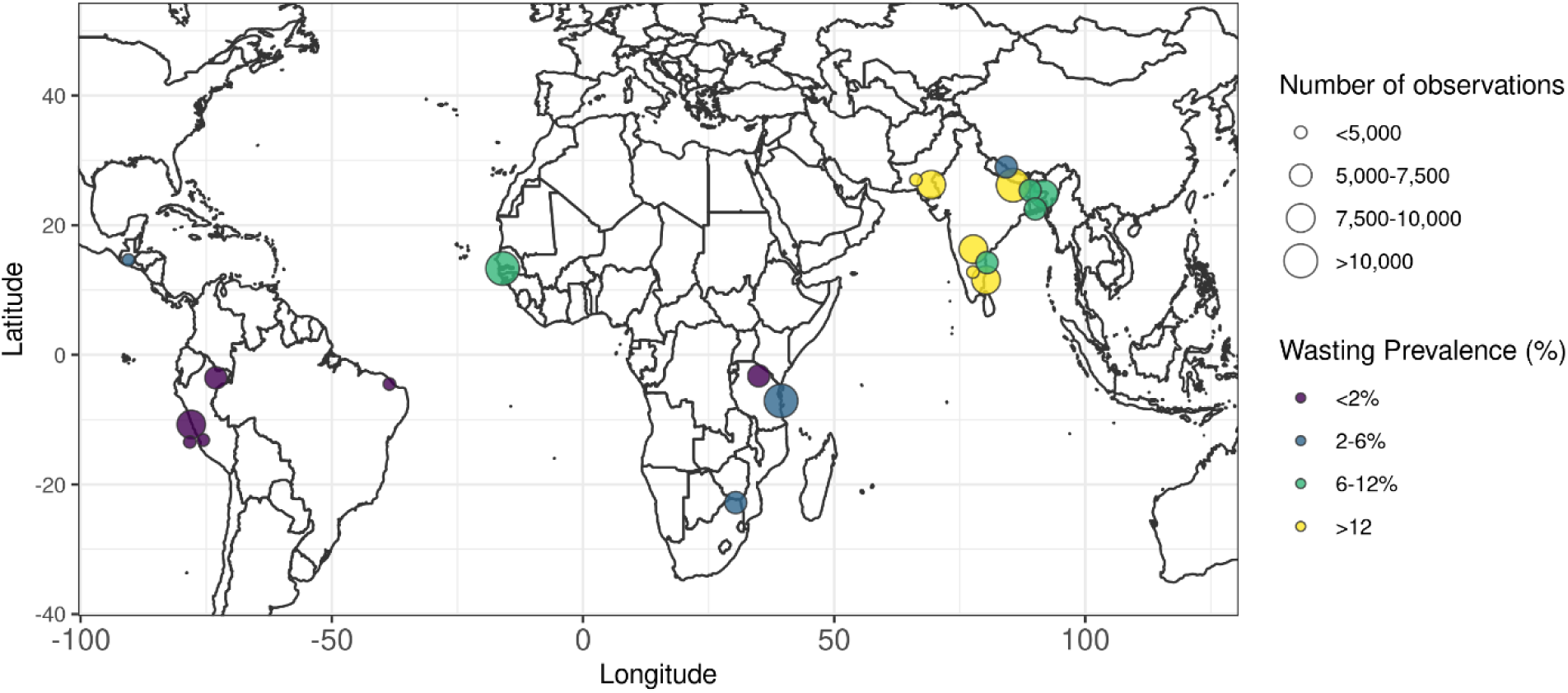
Geographic location of *ki* cohorts. Locations are approximate and jittered slightly for display.

**Extended Data Figure 3.**
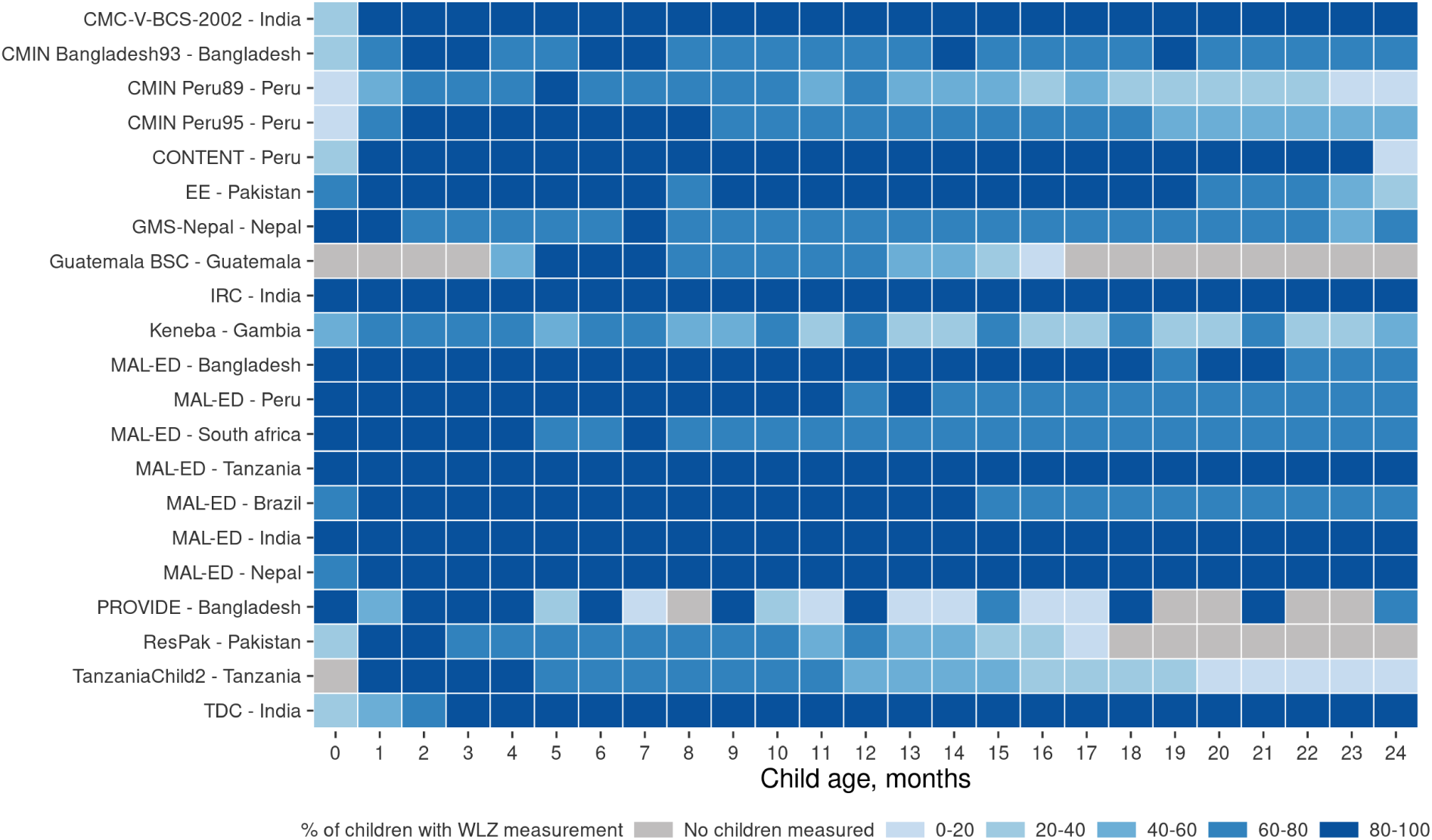
Percentage of enrolled children measured in each ki cohort with monthly measurements.

**Extended Data Figure 4.**
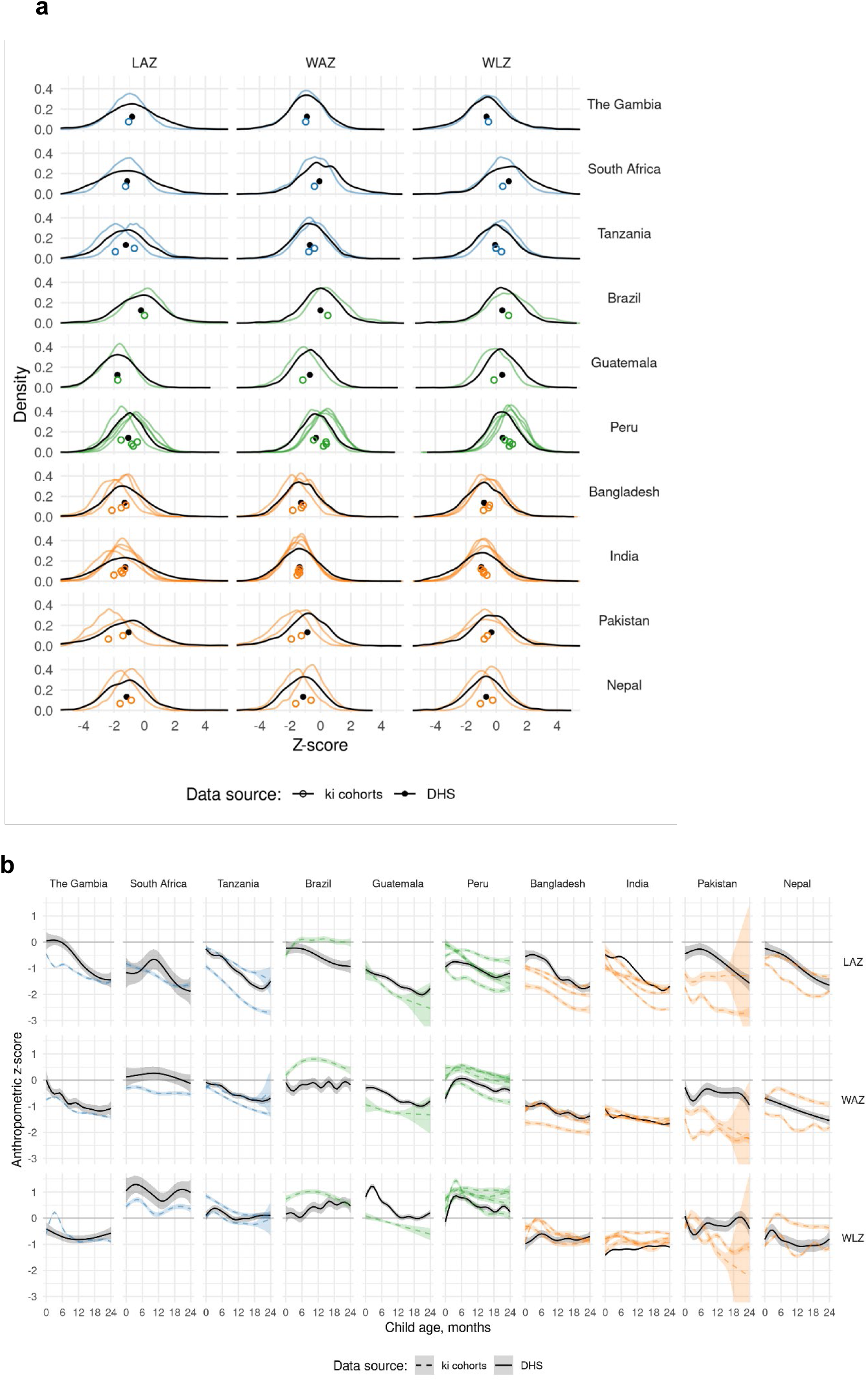
Comparison of cohort anthropometry to population-based samples. **(a)** Kernel density distributions of length-for-age (LAZ), weight-for-age (WAZ), and weight-for-length Z-scores (WLZ) from measurements among children under 24 months old in 21 *ki* longitudinal cohorts (colored line) and among children measured in the most recent population-based, Demographic and Health Survey for each country (black). Sub-Saharan African countries are colored blue, Latin American countries are colored green, and south Asian countries are colored orange. Median Z-scores are denoted with points under the density curves, with open circles for *ki* cohorts and solid points for Demographic and Health Surveys. **(b)** Mean LAZ, WAZ, and WLZ by age and country among 21 *ki* longitudinal cohorts (dashed lines) and in Demographic and Health Surveys (solid). Means estimated with cubic splines and shaded regions show approximate, simultaneous 95% confidence intervals.

**Extended Data Figure 5.**
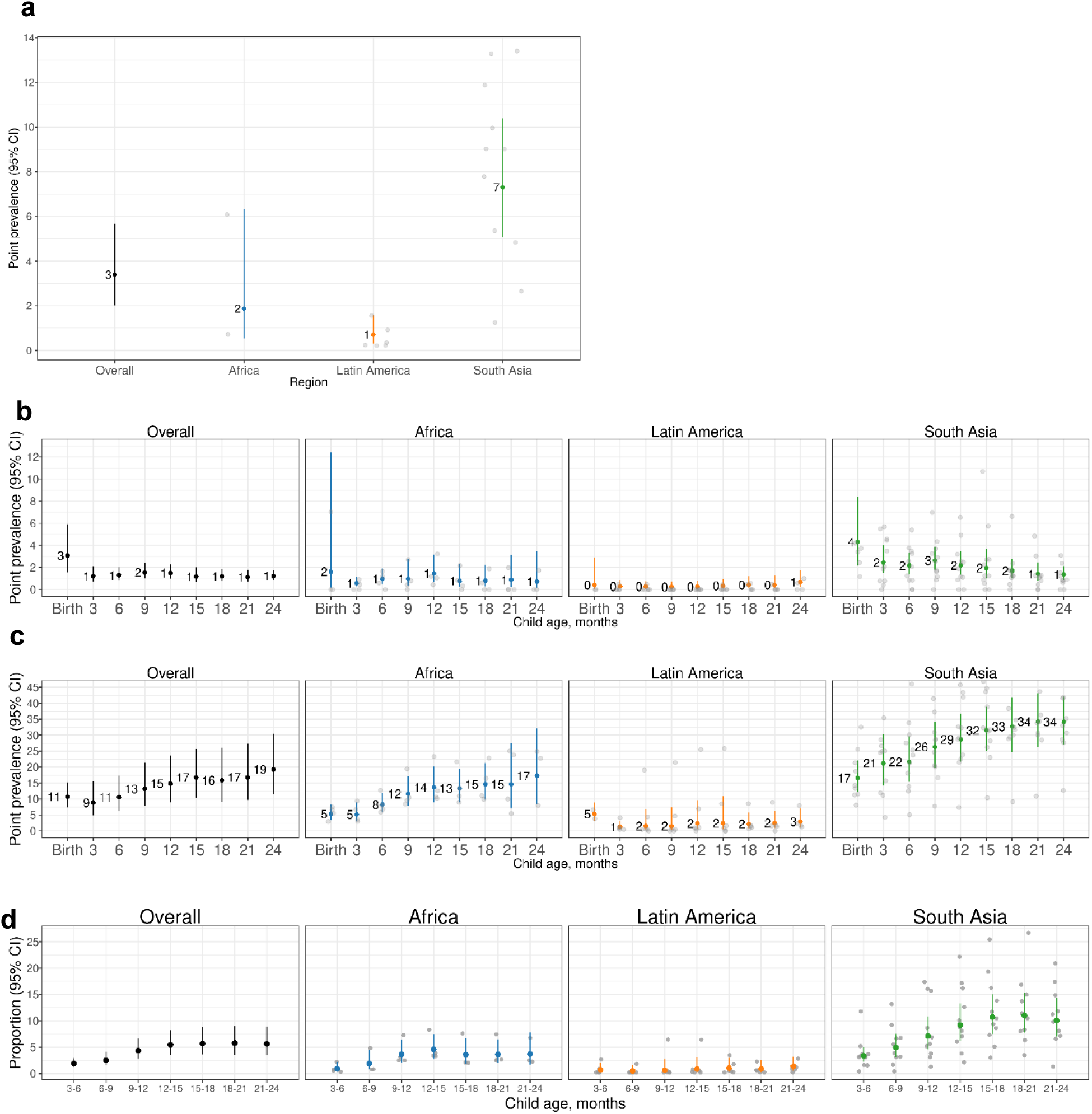
Prevalence of persistent wasting, severe wasting, and underweight by region. **(a)** Proportion of children persistently wasted (≥50% of measurements from birth to 24 months of age, overall and stratified by region. **(b)** Prevalence of severe wasting (WLZ < –3) by age and region. **(c)** Prevalence of underweight (weight-for-age Z-score < –2) by age and region. **(d)** Incidence proportion of concurrent wasting and stunting by age and region. Across all ages before 24 months, 10.6% of children experienced at least one concurrent wasted and stunted measurement.

**Extended Data Figure 6.**
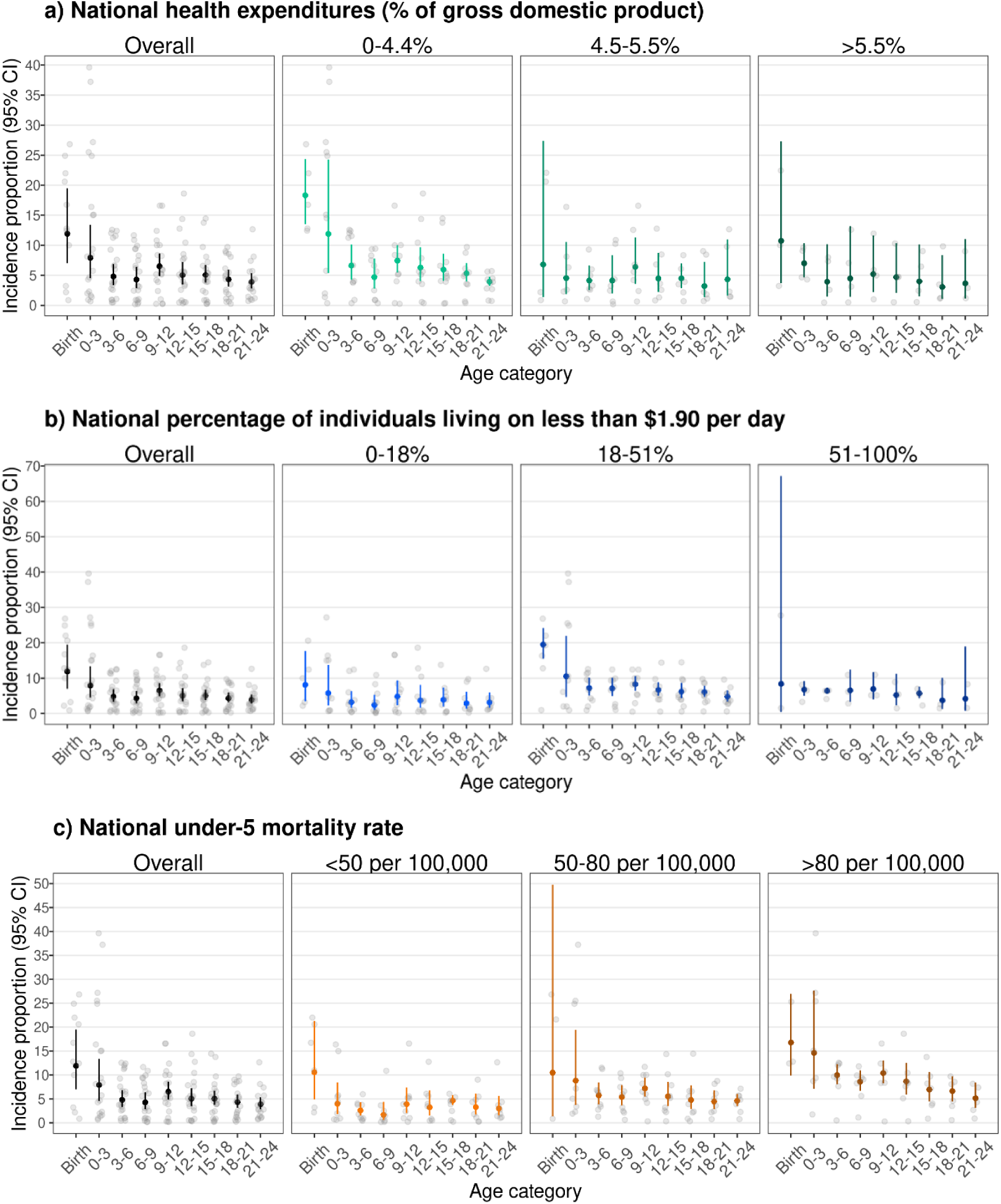
Incidence of wasting by age and country characteristics. Proportion of children at different ages in months who experienced the onset of wasting episodes by **(a)** national health expenditures as a percentage of gross domestic product (0-4.4%: N=5 studies, N=3,305 children; 4.5-5.5%: N=5 studies, N=2,697 children; >5.5%: N=3 studies, N=3,325 children), **(b)** national percentage of individuals living on less than $1.90 US per day (0-18%: N= 5 studies, N= 2,818 children; 18-51%: N= 6 studies, N= 3,584 children; 51-100%: N= 2 studies, N= 3,381 children), and **(c)** national under-5 mortality rate (<50 per 100,000: N= 6 studies, N= 2,743 children; 50-80 per 100,000: N= 3 studies, N= 3,952 children; >80 per 100,000: N= 3 studies, N= 3,611 children). The “Birth: age categories include measurements in the first 7 days of life and the “0-3” age category includes ages 8 days up to 3 months. Vertical bars indicate 95% confidence intervals from random-effects meta-analysis models with restricted maximum likelihood estimation, and gray points indicate cohort-specific estimates.

**Extended Data Figure 7.**
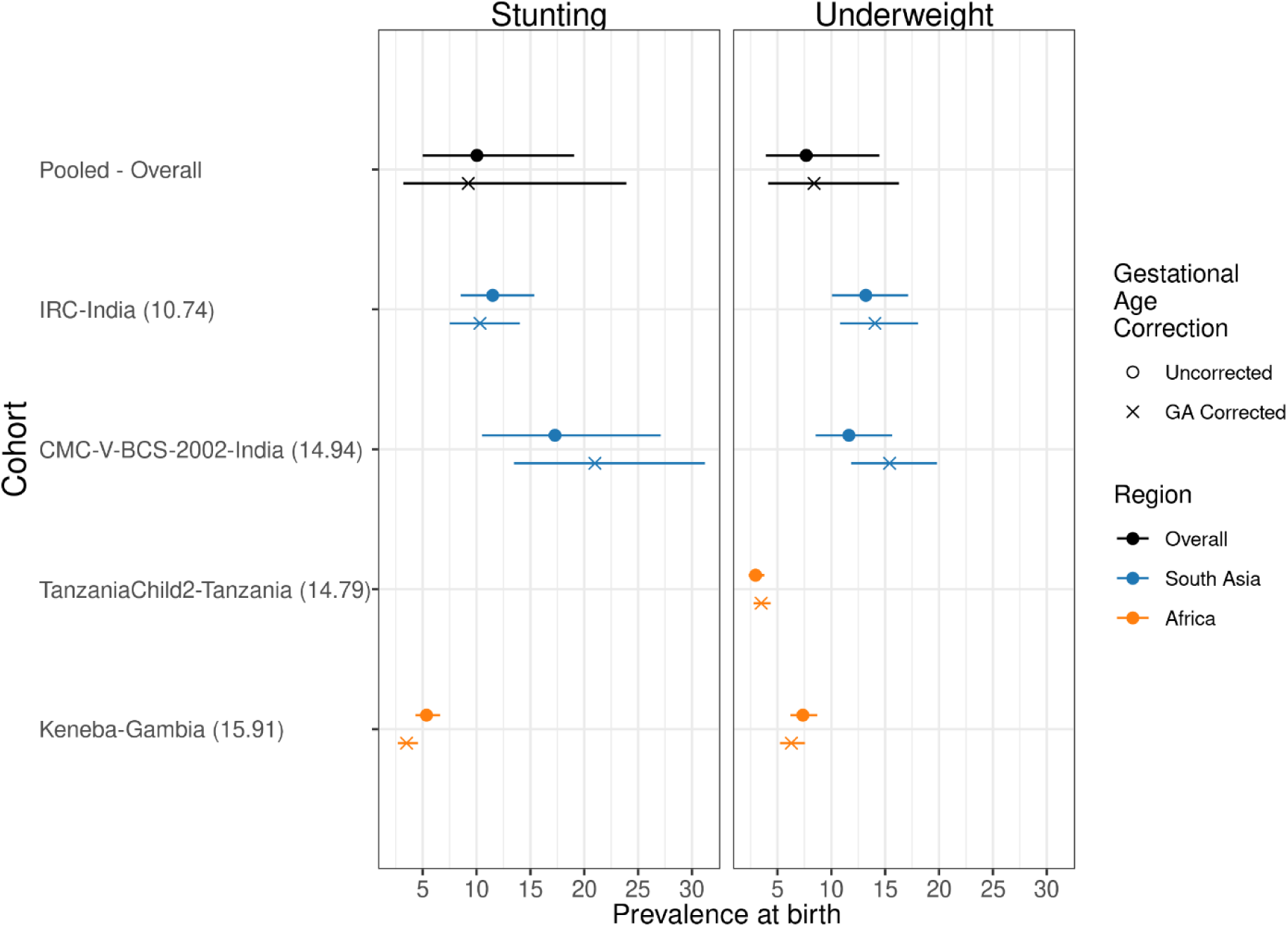
Comparison of underweight and stunting prevalence at birth with and without gestational age correction. This figure includes the results from correcting at-birth Z-scores in the *ki* cohorts that measured gestational age (GA). The corrections are using the Intergrowth standards and are implemented using the R *growthstandards* package (https://ki-tools.github.io/growthstandards/). Overall, the prevalence at birth decreased slightly after correcting for gestational age, but the cohort-specific results are inconsistent. Observations with GA outside of the Intergrowth standards range (<168 or > 300 days) were dropped for both the corrected and uncorrected data. Prevalence increased after GA correction in some cohorts due to high rates of late-term births based on reported GA. There were no length measurements at birth in the Tanzania Child 2 cohort, so they do not have stunting estimates. There were 4,449 measurements used in the underweight analysis and 1,931 measurements used in the stunting analysis. Gestational age was estimates based on mother’s recall of the last menstrual period in the IRC, CMC-V-BCS-2002, and Tanzania Child cohorts, and was based on the Dubowitz method (newborn exam) in the MRC Keneba cohort.

**Extended Data Figure 8.**
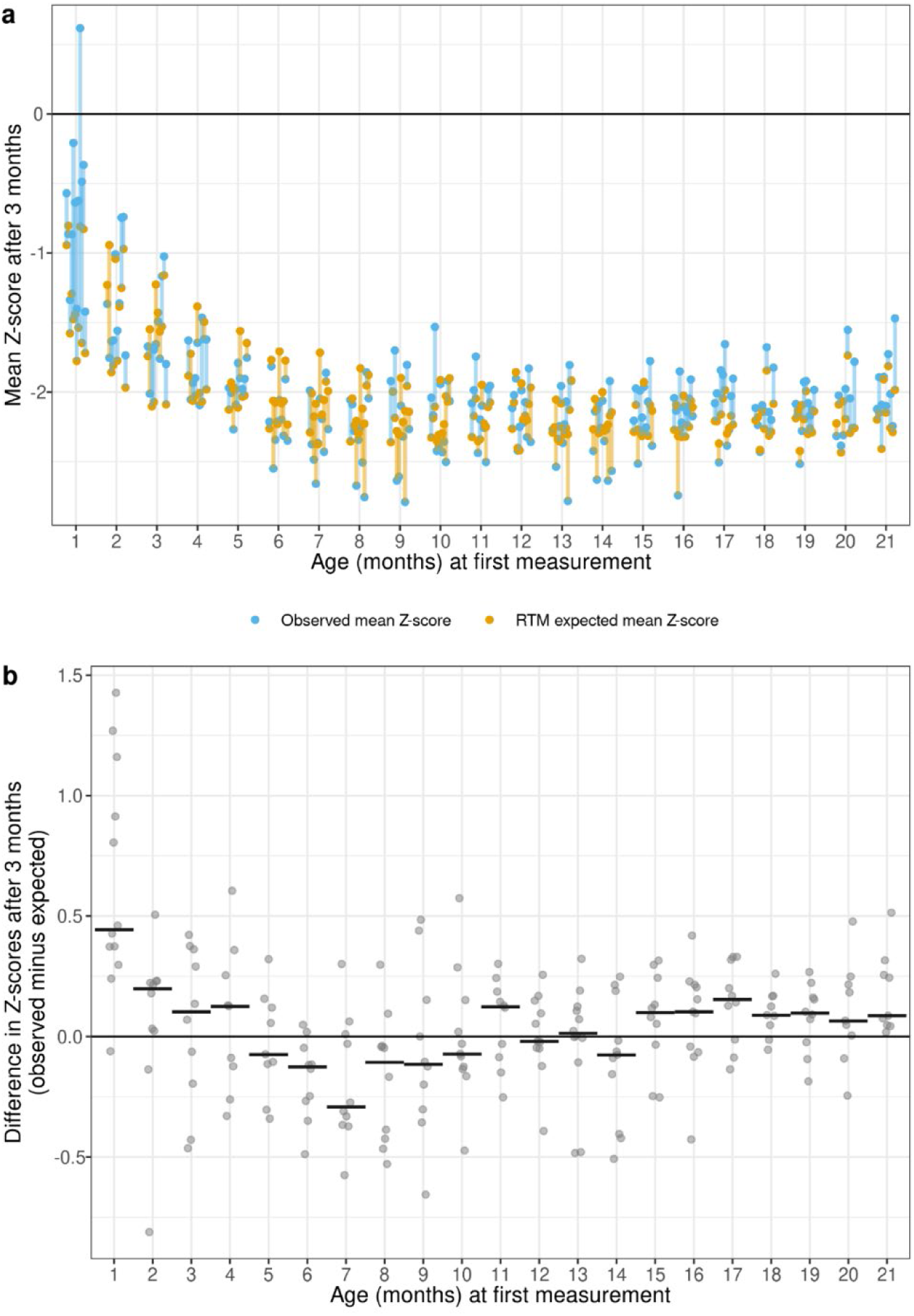
Regression to the mean effects in wasted children. **(a)** Expected mean Z-scores based on the regression to the mean effect (orange) and observed mean Z-scores (blue) 3 months after wasted children are measured. The lines connecting cohort-specific observed and expected WLZ at each age are colored orange if the expected estimate under RTM was higher than the observed mean (indicating lower than expected change in WLZ under RTM alone), and blue if the observed mean was higher than the expected estimate under RTM (indicating higher than expected change in WLZ under RTM alone). For examples, most cohorts experienced larger increases in WLZ than expected in the three-month period beginning in their first month of life (blue lines) and most cohorts experienced smaller increases in WLZ than expected in the three month periods beginning at ages 6-9 months (orange lines). **(b)** Difference between observed means and expected means under a pure RTM effect by cohort, with the median differences by age indicated with horizontal lines. Details on estimation of the RTM effects are in the methods.

**Extended Data Figure 9.**
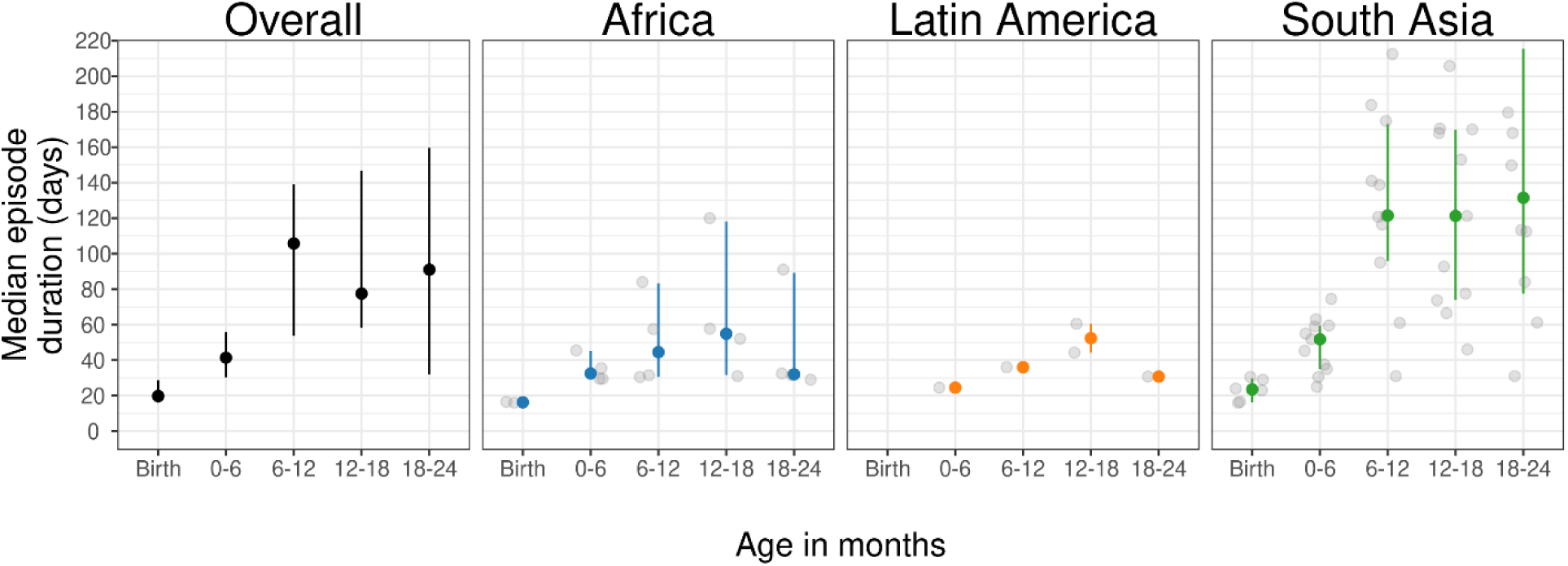
Duration of wasting episodes by child age and region. Duration of wasting episodes that began in each age category, overall (N=787-1,940 episodes per age category) and stratified by region (Africa: 377-916 episodes, Latin America: N= 11-25 episodes, South Asia: N=410-1,146 episodes). The “Birth: age categories includes measurements in the first 7 days of life and the “0-3” age category includes ages 8 days up to 3 months. Estimates were pooled across cohorts using the median of medians method (McGrath et al., 2019).^1^ Vertical bars indicate 95% confidence intervals around the pooled estimates, and gray points indicate cohort-specific estimates. Episodes are assumed to start halfway between non-wasted and wasted measurements, and end halfway between the last wasted measurement and first recovered estimate. Birth episodes start at birth, so episodes at birth are generally shorter that post-birth episodes with the same number of wasted measurements.

**Extended Data Figure 10.**
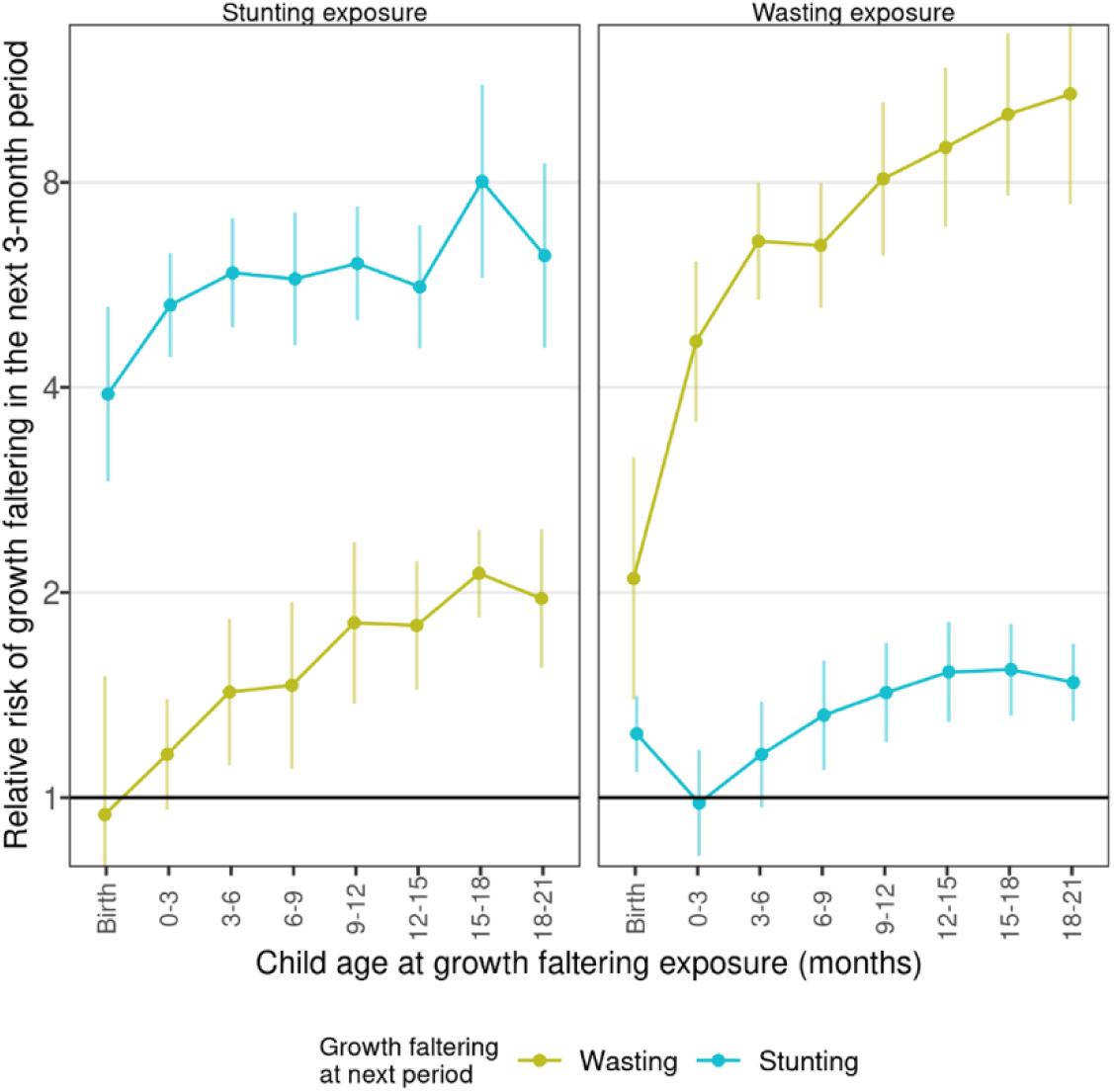
Risk of growth faltering in the subsequent 3 months by whether a child was wasted or stunted at each age. Relative risk of wasting and stunting in the next 3-month period among children who were wasted (left panel) or stunted (right panel) at each age. For example, children who were wasted at birth had an approximately 2-fold increased risk of wasting during the subsequent 0-3 month period, and children who were stunted at birth had an approximately 4-fold increased risk of being stunted during the subsequent 0-3 month period. Points indicate relative risks and vertical lines mark 95% confidence intervals.

**Extended Data Figure 11.**
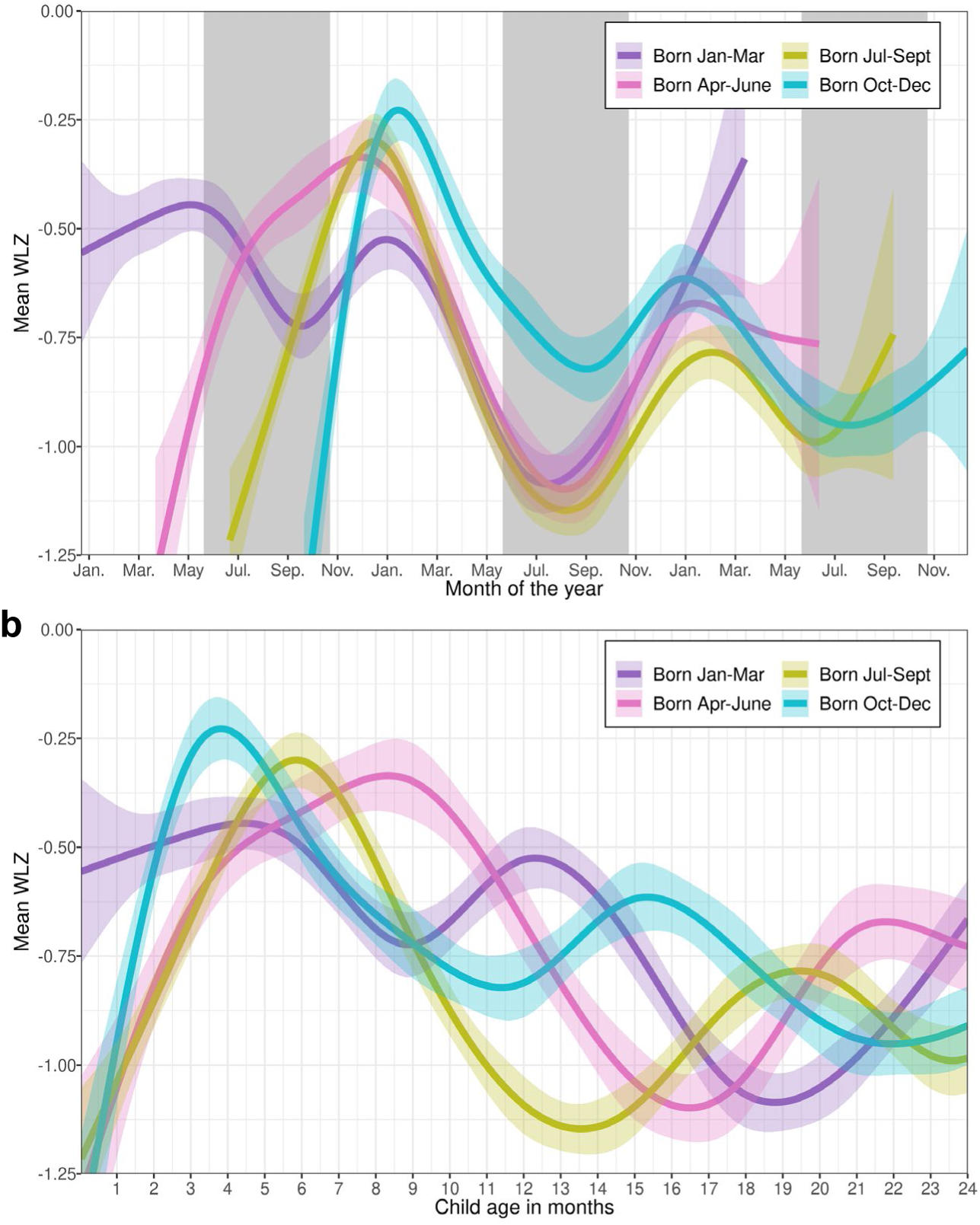
Month of birth affects seasonal patterns in WLZ. **(a)** Mean WLZ by calendar month among South Asian cohorts, with children stratified by birth month. The X-axis begins at January 1^st^ of the year of the first January birthday in each cohort. Grey backgrounds indicate the approximate timing of seasonal monsoons in South Asia (June-September) Shaded regions around spline fits indicate 95% simultaneous confidence intervals. Eleven cohorts, 4,040 children, and 78,573 measurements were used to estimate the splines. South Asian children born in July-September had the lowest mean WLZ overall and children born April-September had larger seasonal declines in WLZ during their second year of life than children born October-March. **(b)** Mean WLZ from birth to age 24 months among children from South Asian cohorts stratified by birth month. Shaded regions around spline fits indicate 95% simultaneous confidence intervals. Data used is the same as in panel (a).

**Extended Data Table 1.**
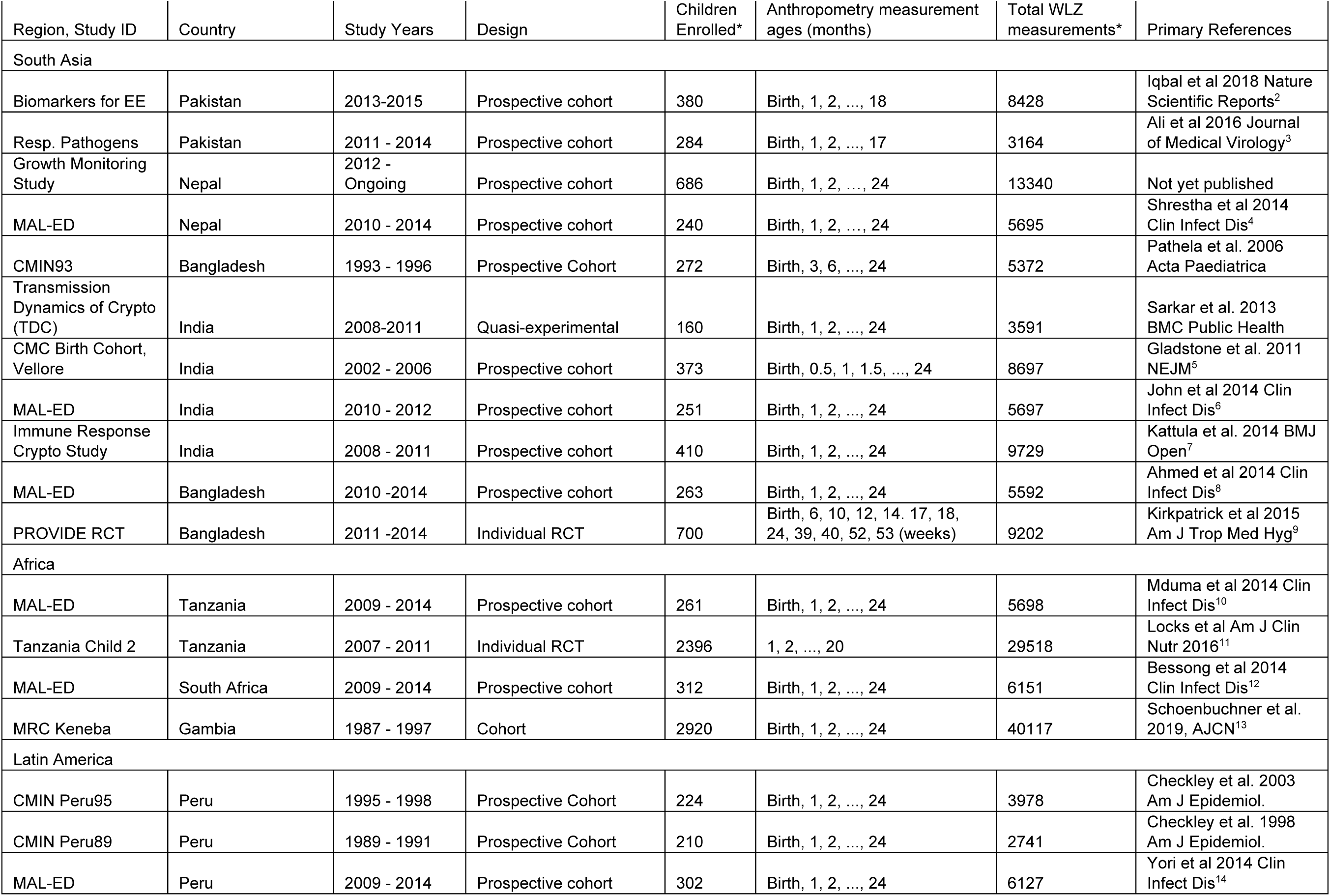
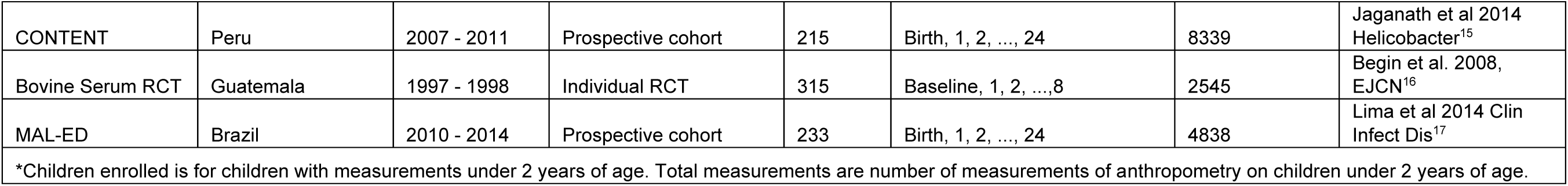
Summary of *ki* cohorts

**Extended Data Table 2.** Summary of baseline characteristics in *ki* cohorts

|  | CMC-V-BCS-2002, India | CMIN<br>Bangladesh93 | CMIN<br>Peru89 | CMIN<br>Peru95 | CONTENT,<br>Peru | EE,<br>Pakistan | GMS-Nepal | Guatemala<br>BSC | IRC, India | Keneba, The<br>Gambia |
| --- | --- | --- | --- | --- | --- | --- | --- | --- | --- | --- |
|  | (N=373) | (N=280) | (N=210) | (N=224) | (N=215) | (N=380) | (N=698) | (N=315) | (N=410) | (N=2954) |
| Sex |  |  |  |  |  |  |  |  |  |  |
| Female | 187 (50.1%) | 122 (43.6%) | 100<br>(47.6%) | 96 (42.9%) | 109 (50.7%) | 185<br>(48.7%) | 328<br>(47.0%) | 162 (51.4%) | 185<br>(45.1%) | 1426 (48.3%) |
| Male | 186 (49.9%) | 158 (56.4%) | 110<br>(52.4%) | 128<br>(57.1%) | 106 (49.3%) | 195<br>(51.3%) | 370<br>(53.0%) | 153 (48.6%) | 225<br>(54.9%) | 1528 (51.7%) |
| Birthweight |  |  |  |  |  |  |  |  |  |  |
| Mean (SD) | 2910 (434) | 2560 (967) | 3550 (492) | 3690 (367) | 3070 (323) | 2640 (506) | 2660<br>(423) |  | 2890<br>(448) | 2970 (421) |
| Maternal age |  |  |  |  |  |  |  |  |  |  |
| Mean (SD) | 24.1 (4.11) |  |  |  |  | 30.0 (3.99) | 24.0<br>(5.09) | 25.2 (6.21) | 23.7<br>(3.68) | 27.4 (7.16) |
| Maternal weight |  |  |  |  |  |  |  |  |  |  |
| Mean (SD) |  |  |  |  |  |  |  |  |  |  |
| Maternal<br>education<br>(years) |  |  |  |  |  |  |  |  |  |  |
| Mean (SD) | 5.43 (4.10) |  |  |  |  | 0.873<br>(2.51) | 2.48<br>(4.01) | 4.07 (2.95) | 5.06<br>(4.61) |  |
| Number of rooms |  |  |  |  |  |  |  |  |  |  |
| 4+ | 14 (3.75%) |  |  |  | 78 (36.3%) |  | 323<br>(46.3%) |  | 17<br>(4.17%) |  |
| 1 | 202 (54.2%) |  |  |  | 44 (20.5%) |  | 49<br>(7.02%) |  | 185<br>(45.3%) |  |
| 2 | 106 (28.4%) |  |  |  | 54 (25.1%) |  | 145<br>(20.8%) |  | 170<br>(41.7%) |  |
| 3 | 51 (13.7%) |  |  |  | 39 (18.1%) |  | 181<br>(25.9%) |  | 36<br>(8.82%) |  |
| Number of<br>children <5yrs |  |  |  |  |  |  |  |  |  |  |
| 1 |  |  |  |  |  |  |  |  | 89<br>(21.7%) |  |
| 2+ |  |  |  |  |  |  |  |  | 321<br>(78.3%) |  |
| Improved<br>sanitation |  |  |  |  |  |  |  |  |  |  |
| 1 |  |  |  |  | 201 (93.5%) |  |  |  |  |  |
| 0 |  |  |  |  | 14 (6.51%) |  |  |  |  |  |
| Food security<br>level |  |  |  |  |  |  |  |  |  |  |
| Food Secure |  |  |  |  |  |  | 479<br>(71.1%) |  |  |  |
| Mildly Food<br>Insecure |  |  |  |  |  |  | 106<br>(15.7%) |  |  |  |
| Food Insecure |  |  |  |  |  |  | 89<br>(13.2%) |  |  |  |

|  | MAL-ED,<br>Bangladesh | MAL-ED,<br>Brazil | MAL-ED,<br>India | MAL-ED,<br>Nepal | MAL-ED,<br>Peru | MAL-ED,<br>South Africa | MAL-ED,<br>Tanzania | PROVIDE,<br>Bangladesh | ResPak,<br>Pakistan | Tanzania<br>Child2 | TDC,<br>India |
| --- | --- | --- | --- | --- | --- | --- | --- | --- | --- | --- | --- |
|  | (N=265) | (N=233) | (N=251) | (N=240) | (N=303) | (N=314) | (N=262) | (N=700) | (N=284) | (N=2400) | (N=160) |
| Sex |  |  |  |  |  |  |  |  |  |  |  |
| Female | 136 (51.3%) | 113<br>(48.5%) | 138<br>(55.0%) | 110<br>(45.8%) | 143<br>(47.2%) | 159 (50.6%) | 133 (50.8%) | 332 (47.4%) | 136<br>(47.9%) | 1184<br>(49.3%) | 75<br>(46.9%) |
| Male | 129 (48.7%) | 120<br>(51.5%) | 113<br>(45.0%) | 130<br>(54.2%) | 160<br>(52.8%) | 155 (49.4%) | 129 (49.2%) | 368 (52.6%) | 148<br>(52.1%) | 1216<br>(50.7%) | 85<br>(53.1%) |
| Birthweight |  |  |  |  |  |  |  |  |  |  |  |
| Mean (SD) | 2800 (412) | 3340<br>(483) | 2890<br>(442) | 2980 (390) | 3130<br>(430) | 3130 (464) | 3180 (452) | 2780 (371) | 2930 (525) | 3230 (474) | 2910<br>(450) |
| Maternal age |  |  |  |  |  |  |  |  |  |  |  |
| Mean (SD) | 24.8 (4.99) | 24.8<br>(5.53) | 23.9<br>(4.10) | 26.4 (3.76) | 24.2<br>(6.06) | 26.4 (6.86) | 28.4 (6.67) | 24.7 (4.65) |  | 26.4 (5.04) |  |
| Maternal weight |  |  |  |  |  |  |  |  |  |  |  |
| Mean (SD) | 49.6 (8.54) | 61.9<br>(11.8) | 50.4<br>(9.37) | 56.3 (8.27) | 55.5<br>(8.95) | 67.5 (14.9) | 55.7 (9.11) | 49.3 (9.41) |  | 62.2 (11.7) |  |
| Maternal education (years) |  |  |  |  |  |  |  |  |  |  |  |
| Mean (SD) | 5.63 (2.58) | 9.18<br>(2.81) | 7.88<br>(3.17) | 8.77 (3.44) | 7.81<br>(2.79) | 10.3 (1.94) | 6.17 (1.79) | 4.33 (3.62) |  | 7.90 (2.27) |  |
| Number of rooms |  |  |  |  |  |  |  |  |  |  |  |
| 4+ | 12 (4.96%) | 127<br>(60.5%) | 25<br>(10.6%) | 131<br>(55.5%) | 139<br>(51.1%) | 196 (76.3%) | 108 (43.2%) | 23 (3.29%) |  |  | 5<br>(3.13%) |
| 1 | 152 (62.8%) | 4 (1.90%) | 84<br>(35.7%) | 52 (22.0%) | 19<br>(6.99%) | 14 (5.45%) | 13 (5.20%) | 507 (72.4%) |  |  | 91<br>(56.9%) |
| 2 | 50 (20.7%) | 20<br>(9.52%) | 78<br>(33.2%) | 31 (13.1%) | 52<br>(19.1%) | 22 (8.56%) | 63 (25.2%) | 108 (15.4%) |  |  | 49<br>(30.6%) |
| 3 | 28 (11.6%) | 59<br>(28.1%) | 48<br>(20.4%) | 22 (9.32%) | 62<br>(22.8%) | 25 (9.73%) | 66 (26.4%) | 62 (8.86%) |  |  | 15<br>(9.38%) |
| Number of children <5yrs |  |  |  |  |  |  |  |  |  |  |  |
| 1 |  |  |  |  |  |  |  | 512 (73.1%) |  | 1640<br>(68.6%) |  |
| 2+ |  |  |  |  |  |  |  | 188 (26.9%) |  | 749<br>(31.4%) |  |
| Improved sanitation |  |  |  |  |  |  |  |  |  |  |  |
| 1 | 204 (84.3%) | 206<br>(98.1%) | 108<br>(46.4%) | 235<br>(99.6%) | 65<br>(24.7%) | 4 (1.60%) |  | 58 (96.7%) |  |  |  |
| 0 | 38 (15.7%) | 4 (1.90%) | 125<br>(53.6%) | 1 (0.424%) | 198<br>(75.3%) | 246 (98.4%) |  | 2 (3.33%) |  |  |  |
| Food security<br>level |  |  |  |  |  |  |  |  |  |  |  |
| Food Secure | 161 (83.0%) | 3 (2.33%) | 190<br>(89.6%) | 94 (73.4%) | 27<br>(23.9%) | 132 (56.7%) |  |  |  |  |  |
| Mildly Food<br>Insecure | 4 (2.06%) | 11<br>(8.53%) | 5 (2.36%) | 15 (11.7%) | 29<br>(25.7%) | 19 (8.15%) |  |  |  |  |  |
| Food Insecure | 29 (14.9%) | 115<br>(89.1%) | 17<br>(8.02%) | 19 (14.8%) | 57<br>(50.4%) | 82 (35.2%) |  |  |  |  |  |

## Materials and Methods

### Study designs and inclusion criteria

We included all longitudinal observational studies and randomized trials available through the ki project on April 2018 that met five inclusion criteria (Extended Data Figure 1): 1) conducted in low- or middle-income countries; 2) enrolled children between birth and age 24 months and measured their length and weight repeatedly over time; 3) did not restrict enrollment to acutely ill children; 4) enrolled at least 200 children; 5) collected anthropometry measurements at least monthly. The frequency of measurements was assessed by calculating the median days between measurements. Our pre-specified analysis protocol stipulated that if randomized trials found effects of interventions on growth, the analysis would only include the control arm only; however, all interventional trials that met the inclusion criteria had null effects on growth, so all arms were included. We included all children under 24 months of age, assuming months were 30.4167 days. We excluded extreme measurements of WLZ > 5 or <–5 and of WAZ <-6 or > 5, consistent with 2006 WHO Growth Standards recommendations.^1^ We checked for cohort-wide anthropometry measurement quality by plotting Z-score densities and calculating the proportion of length measurements where length decreased beyond the expected technical error of measurement compared to the last measurement on a child. One cohort, MAL-ED Pakistan, was excluded because measurements exhibited a multimodal WLZ distribution with scores binned at –2, – 1.5, and –1 instead of continuously distributed. Some young infants with weight and height measured were missing WLZ, as WLZ cannot be calculated when child length is below 45cm (0.8% of measurements before age 1 month, with a mean WAZ for those children of -2.9 z, and 10.2% of children measured at birth), so using WLZ underestimates wasting at birth.^2^

### Outcome definitions

We used the following outcome measures in the analysis:

<u>Weight-for-Length Z-scores</u> were calculated using the 2006 WHO growth standards,3 and mean WLZ was calculated within strata of interest. We used the medians of triplicate measurements of lengths and weights of children from pre-2006 cohorts to re-calculate Z scores to the 2006 standard.

<u>Prevalent wasting</u> was defined as the proportion of measurements within a specific stratum (e.g., age) below the 2006 WHO standard –2 WLZ and prevalent severe wasting was analogously below the 2006 WHO standard –3 WLZ. For each age, we included children with WLZ measurements within one month before and after that age in the point prevalence estimate (e.g., for point prevalence at 6 months, we include children aged 5-7 months).

<u>Incident wasting episodes</u> were defined as a change in WLZ from above –2 Z in the prior measurement to below –2 Z in the current measurement. Similarly, we defined severe wasting episodes using –3 Z cutoff. We assumed a 60-day washout period between episodes of wasting before a new episode of wasting could occur (Fig 4a). Children were considered at risk for wasting at birth, so children born wasted were considered to have an incident episode of wasting at birth. Children were also assumed to be at risk of wasting at the first measurement in studies that enrolled children after birth; thus, children wasted at the first measurement in a non-birth cohort were assumed to have incident wasting occurring at the age halfway between birth and the first measurement.

<u>Incidence proportion</u> of wasting was calculated during a defined age range (i.e., 6-12 months) as the proportion of children not wasted at the start of the period who became wasted during the age period (the proportion of children who had the onset of new episodes during the period). This differs from period prevalence, which would include any children who had any measurement of WLZ <–2 during the period (including children who began the period wasted). Period prevalence would thus additionally include in the numerator and denominator children who were wasted at the start of the period, whereas incidence proportion excludes them.

<u>Recovery from wasting</u> was defined as a WLZ change from below to above –2 Z among children who are currently wasted or severely wasted. We required a child to maintain WLZ above –2 for 60 days to be considered “recovered”. Children were only considered “at risk” for recovery if their prior measurement was below –2 WLZ. We measured the proportion of children who recover from moderate wasting (WLZ <–2) within 30, 60, and 90 days of the onset of the episode. We assumed recovery from wasting was spontaneous because we did not have consistent information on referral guidelines across cohorts in the analysis, but some children with moderate wasting may have been referred to clinical facilities for outpatient treatment. Wasting recovery may thus be higher in these highly monitored cohorts than in the general population due to treatment referrals.

<u>Wasting duration</u> was estimated by counting the days between the onset of wasting and recovery within an individual child. We assumed that the episode started or ended at the midpoint between measurements. For example, if a child was not wasted at age 40 days, wasted at age 70 days, and not wasted at age 100 days, the duration of the wasting episode was: (70-40)/2 + (100-70)/2 = 30 days. We calculated the median duration of wasting episodes among cohorts where children recovered during the study period from >50% of observed wasting episodes, as the duration of episodes that extended beyond study period is unknown. Therefore, the median duration could only be calculated when less than half of episodes were censored.

<u>Incidence rate</u> of wasting was calculated during defined age ranges as the number of incident episodes of wasting per 1,000 child-days at risk during the age range. Children were considered “at risk” for incident wasting episodes if they were not currently wasted and were classified as recovered from any prior wasting episode (beyond the 60-day washout period). Therefore, wasted children, or children within the washout period, did not contribute to the person-time at risk for wasting used to calculate wasting incidence. To calculate person-time at risk of wasting, we assumed that the onset of a wasting episode occurred when the child’s age was at the midpoint between the measurement of WLZ ≥–2 and the measurement of WLZ <–2, and conversely the time of recovery occurred when the child’s age was at the midpoint between the last measurement of WLZ <–2 and the first measurement of WLZ ≥–2 within the 60-day washout period (Fig 4a).

<u>Persistent wasting</u> was defined as a >50% longitudinal prevalence of below –2 WLZ (wasting), and analogously >50% longitudinal prevalence of below –3 WLZ for persistent severe wasting.^4^

<u>Concurrent prevalence of wasting and stunting</u> was defined as the proportion of measurements at a specific age when a child was both wasted and stunted at the same measurement. For each age, we included children with WLZ and LAZ measurements within one month before and after that age in the point prevalence estimate (e.g., for point prevalence at 6 months, we include children aged 5-7 months).

<u>Prevalent underweight</u> was defined as the proportion of measurements at a specific age below the 2006 WHO standard –2 WAZ. For each age, we included children with WAZ measurements within one month before and after that age in the point prevalence estimate (e.g., for point prevalence at 6 months, we include children aged 5-7 months).

### Subgroups of interest

We stratified the above outcomes of interest within the following subgroups: Child age, grouped into one-, three-, or six-month intervals, (depending on the outcome); the region of the world (Asia, sub-Saharan Africa, Latin America); the month of the year, and the combinations of the above categories. We also pooled wasting incidence proportion by categories of three country-level characteristics: the percentage of gross domestic product devoted to healthcare goods and spending (obtained from the United Nations Development Programme),^5^ the percentage of the country living on less than $1.90 US per day, and under-5 mortality rates (both obtained from the World Bank).^6^ Within each country, in years without available data, we linearly interpolated values from the nearest years with available data and extrapolated values within 5 years of available data using linear regression models based on all available years of data.

## Statistical analysis

All analyses were conducted in R version 4.0.5. Supplementary pooled, regional, and cohort-specific results, and sensitivity analyses are available in online supplements at (https://child-growth.github.io/wasting).

### Fixed and random effects models

We conducted a 2-stage individual participant meta-analysis, estimating wasting outcomes within specific cohorts and pooling estimates within each age strata using random effects models. For example, we estimated each age-specific mean using a two-step process. We first estimated the mean in each cohort, and then pooled age-specific means across cohorts allowing for a cohort-level random effect. We estimated overall pooled effects, and pooled estimates specific to South Asian, African, or Latin American cohorts, depending on the analysis. We repeated the pooling of all statistics presented in the figures using fixed effects models as a sensitivity analysis (https://child-growth.github.io/wasting/fixed-effects.html). The pooling methods are described in greater detail in Benjamin-Chung (2020).^7^ All pooling except for episode duration was completed using the rma() function from the “metafor” package in the R language (version 3.4.0).^8^ Median durations were pooled across cohorts using the unweighted median of medians method (McGrath et al., 2019).^9^

### Fitted spline curves

Fitted smoothers used in manuscript figures were fit using cubic splines and generalized cross-validation.10 We estimated approximate 95% simultaneous confidence intervals around the cubic splines using a parametric bootstrap that resampled from the posterior the generalized additive model parameter variance-covariance matrix.11

### Seasonality analysis

We compared mean WLZ over day of the year, over child birth day, and over seasons defined by rainfall. We estimated mean WLZ by cohort over day of the year and child birth day using cubic splines (Fig. 3a, c).^12^ Splines of WLZ over day of the year were plotted over monthly mean rainfall averaged over the years a study measured child anthropometry below ages 24 months (Fig. 3a). We pulled monthly precipitation values from Terrraclimate, a dataset that combines readings from WorldClim data, CRU Ts4.0, and the Japanese 55-year Reanalysis Project.^13^ For each study region, we averaged all readings within a 50 km radius from the study coordinates. If GPS locations were not in the data for a cohort, we used the approximate location of the cohort based on the published descriptions of the cohort. Monthly measurements were matched to study data based on the calendar month and year in which measurements were taken.

The season of peak rainfall was defined as the three-month period with the highest mean rainfall. Mean differences in WLZ between three-month quarters was estimated using linear regression models. We compared the consecutive three months of the maximum average rainfall over the study period, as well as the three months prior and the three months after the maximum-rainfall period, to a reference level of the three months opposite the calendar year of the maximum-rainfall period. We used all WLZ measurements of children under two years of age (e.g., if June-August was the period of maximum rainfall, the reference level is child mean WLZ during January-March).

Estimates were unadjusted for other covariates because we assumed that seasonal effects on WLZ were exogenous and could not be confounded. Mean differences in WLZ were pooled across cohorts using random-effects models, with cohorts grouped by the Walsh and Lawler seasonality index.^14^ Cohorts from years with a seasonal index ≥0.9 were classified as occurring in locations with high seasonality, cohorts with a seasonal index <0.9 and ≥0.7 were classified as occurring in locations with medium seasonality, and cohorts with a seasonal index <0.7 were classified as occurring in locations with low seasonality.

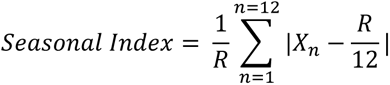

Where R = total annual precipitation and *X_n_* = monthly precipitation.

#### Estimation of mean LAZ, WAZ, and WLZ by age in Demographic and Health Surveys and *ki* cohorts

We downloaded standard DHS individual recode files for each country from the DHS program website (https://dhsprogram.com/). We used the most recent standard DHS datasets for the individual women’s, household, and length and weight datasets from each country, and we estimated age-stratified mean LAZ, WAZ, and WLZ from ages 0 to 24 months within each DHS survey, accounting for the complex survey design and sampling weights. See Benjamin-Chung et. al (2020) for additional details on the DHS data cleaning and analysis.^7^ We compared DHS estimates with mean LAZ, WAZ, and WLZ by age in the *ki* study cohorts with penalized cubic-splines with bandwidths chosen using generalized cross-validation.^12^ We did not seasonally adjust DHS measurements.

#### Sensitivity analyses

We estimated incidence rates and of wasting after excluding children born or enrolled wasted (Fig. 4b). The rationale for this sensitivity analysis is that incident cases at birth imply a different type of intervention (i.e., prenatal) compared with postnatal onset of wasting. We estimated the overall and region stratified prevalence of persistent wasting and of underweight and severe wasting by age (Extended data fig. 5). Within cohorts that measured child gestational age at birth, we estimated the prevalence of stunting and underweight at birth both uncorrected and corrected for gestational age at birth using the Intergrowth standards (Extended data figure 6).^15^ We also assessed how much change in Z-scores after children became wasted could be explained by regression to the mean (RTM) and how much is catch-up growth beyond that expected by RTM.^16^ We calculated the cohort-specific RTM effect and plotted the mean Z-scores expected from RTM 3 months later among children who were wasted at each age in months from birth to 21 months. We used the cohort means as the population mean Z-scores, and compared the expected mean WLZ with the observed mean WLZ (Extended Data Fig. 8).^17, 18^ We also compared wasting prevalence defined using middle-upper-arm circumference (MUAC), an alternative measurement for classifying wasting, with wasting prevalence estimated using WLZ within the cohort that measured MUAC (https://child-growth.github.io/wasting/muac.html). We also re-estimate primary results dropping observations of children at birth within the MRC Keneba cohort, which used a different team to measure child anthropometry at birth from the trained anthropometrists used in follow-up measurements (https://child-growth.github.io/wasting/no-kenaba.html). We also examined the effect of shorter (30 day) and longer (90) day washout period when determining if a child was again at risk when estimating wasting incidence and wasting recovery rates (https://child-growth.github.io/wasting/ir_sensitivity.html). Lastly, we compared estimates pooled using random effects models, which are more conservative in the presence of study heterogeneity, with estimates pooled using fixed effects (inverse variance weighted) models (https://child-growth.github.io/wasting/fixed-effects.html).

## Data and code availability

The data that support the findings of this study are available from the Bill and Melinda Gates Foundation Knowledge Integration project upon reasonable request. Replication scripts for this analysis are available here: https://osf.io/9xyqv/.

## Acknowledgments

This research was financially supported by a global development grant (OPP1165144) from the Bill & Melinda Gates Foundation to the University of California, Berkeley, CA, USA. We would also like to thank the following collaborators on the included cohorts and trials for their contributions to study planning, data collection, and analysis: Muhammad Sharif, Sajjad Kerio, Ms. Urosa, Ms. Alveen, Shahneel Hussain, Vikas Paudel (Mother and Infant Research Activities), Anthony Costello (University College London), Benjamin Torun, Lindsey M Locks, Christine M McDonald, Roland Kupka, Ronald J Bosch, Rodrick Kisenge, Said Aboud, Molin Wang, and all other members of the study staffs and field teams. We would also like to thank all study participants and their families for their important contributions.

## Author contributions

Conceptualization: A.M., J.B., J.M.C., K.H.B., P.C., B.F.A

Funding Acquisition: J.M.C., A.E.H., M.J.V., B.F.A.

Data curation: A.M., J.B., J.C., O.S., W.C., A.N., N.N.P., W.J., E.J., E.O.C., S.D., N.H., I.M., H.L., R.H., V.S., J.H., T.N.

Formal analyses: A.M., J.B., J.C., O.S., W.C., A.N., N.N.P., W.J., E.J., E.O.C., S.D., N.H., I.M., H.L., V.S., B.F.A

Methodology: A.M., J.B., J.M.C, J.C., O.S., N.H., I.M., A.E.H., M.J.V.,K.H.B., P.C., B.F.A.

Visualization: A.M.,J.B., A.N., N.N.P., S.D., A.S., J.C., R.H., E.J., K.H.B., P.C., B.F.A.

Writing – Original Draft Preparation: A.M., J.B., B.F.A.

Writing – Review & Editing: A.M., J.B., J.M.C., K.H.B., P.C., B.F.A., ki Child Growth Consortium members

## Additional information

Supplementary Information is available for this paper at https://child-growth.github.io/wasting.

Correspondence and requests for materials should be addressed to Andrew Mertens and Benjamin F. Arnold.

## References

1. WHO | UNICEF-WHO-The World Bank: Joint child malnutrition estimates - Levels and trends. WHO http://www.who.int/nutgrowthdb/estimates/en/.

2. Onis, M. de. WHO Child Growth Standards based on length/height, weight and age. Acta Paediatr. 95, 76–85 (2006).

3. Rytter, M. J. H., Kolte, L., Briend, A., Friis, H. & Christensen, V. B. The Immune System in Children with Malnutrition—A Systematic Review. PLOS ONE 9, e105017 (2014).

4. Chang, C. Y. et al. Children successfully treated for moderate acute malnutrition remain at risk for malnutrition and death in the subsequent year after recovery. J. Nutr. 143, 215–220 (2013).

5. McDonald, C. M. et al. The effect of multiple anthropometric deficits on child mortality: meta-analysis of individual data in 10 prospective studies from developing countries. Am. J. Clin. Nutr. 97, 896–901 (2013).

6. Schoenbuchner, S. M. et al. The relationship between wasting and stunting: a retrospective cohort analysis of longitudinal data in Gambian children from 1976 to 2016. Am. J. Clin. Nutr. doi:10.1093/ajcn/nqy326.

7. Khandelwal, N. et al. Determinants of motor, language, cognitive, and global developmental delay in children with complicated severe acute malnutrition at the time of discharge: An observational study from Central India. PLOS ONE 15, e0233949 (2020).

8. Sudfeld, C. R. et al. Malnutrition and Its Determinants Are Associated with Suboptimal Cognitive, Communication, and Motor Development in Tanzanian Children. J. Nutr. 145, 2705–2714 (2015).

9. Keats, E. C. et al. Effective interventions to address maternal and child malnutrition: an update of the evidence. *Lancet Child Adolesc*. Health 5, 367–384 (2021).

10. Stobaugh, H. C. et al. Relapse after severe acute malnutrition: A systematic literature review and secondary data analysis. Matern. Child. Nutr. 15, e12702 (2019).

11. O’Sullivan, N. P., Lelijveld, N., Rutishauser-Perera, A., Kerac, M. & James, P. Follow-up between 6 and 24 months after discharge from treatment for severe acute malnutrition in children aged 6-59 months: A systematic review. PLOS ONE 13, e0202053 (2018).

12. Rana, R., McGrath, M., Sharma, E., Gupta, P. & Kerac, M. Effectiveness of Breastfeeding Support Packages in Low- and Middle-Income Countries for Infants under Six Months: A Systematic Review. Nutrients 13, 681 (2021).

13. Dewey, K. G. et al. Preventive small-quantity lipid-based nutrient supplements reduce severe wasting and severe stunting among young children: an individual participant data meta-analysis of randomized controlled trials. Am. J. Clin. Nutr. nqac232 (2022) doi:10.1093/ajcn/nqac232.

14. Lenters, L., Wazny, K. & Bhutta, Z. A. Management of Severe and Moderate Acute Malnutrition in Children. in Reproductive, Maternal, Newborn, and Child Health: Disease Control Priorities, Third Edition *(Volume 2)* (eds. Black, R. E., Laxminarayan, R., Temmerman, M. & Walker, N.) (The International Bank for Reconstruction and Development / The World Bank, 2016).

15. Das, J. K., Salam, R. A., Imdad, A. & Bhutta, Z. A. Infant and Young Child Growth. in Reproductive, Maternal, Newborn, and Child Health: Disease Control Priorities, Third Edition (Volume 2) (eds. Black, R. E., Laxminarayan, R., Temmerman, M. & Walker, N.) (The International Bank for Reconstruction and Development / The World Bank, 2016).

16. Bhutta, Z. A. et al. What works? Interventions for maternal and child undernutrition and survival. Lancet 371, 417–440 (2008).

17. Nabwera, H. M., Fulford, A. J., Moore, S. E. & Prentice, A. M. Growth faltering in rural Gambian children after four decades of interventions: a retrospective cohort study. Lancet Glob. Health 5, e208–e216 (2017).

18. Benjamin-Chung, J. et. al. (submitted). Early childhood linear growth faltering in low-and middle-income countries. (2020).

19. Stobaugh, H. C. et al. Children with Poor Linear Growth Are at Risk for Repeated Relapse to Wasting after Recovery from Moderate Acute Malnutrition. J. Nutr. 148, 974–979 (2018).

20. Osgood-Zimmerman, A. et al. Mapping child growth failure in Africa between 2000 and 2015. Nature 555, 41–47 (2018).

21. Bulti, A. et al. Improving estimates of the burden of severe acute malnutrition and predictions of caseload for programs treating severe acute malnutrition: experiences from Nigeria. Arch. Public Health 75, (2017).

22. Egata, G., Berhane, Y. & Worku, A. Seasonal variation in the prevalence of acute undernutrition among children under five years of age in east rural Ethiopia: a longitudinal study. BMC Public Health 13, (2013).

23. Kinyoki, D. K. et al. Space-time mapping of wasting among children under the age of five years in Somalia from 2007 to 2010. Spat. Spatio-Temporal Epidemiol. 16, 77–87 (2016).

24. Peppard, T., et al. (submitted). Combined longitudinal growth cohorts from infants born in South Asia, Sub-Saharan Africa and Latin America. (2020).

25. WHO Multicentre Growth Reference Study Group. WHO child growth standards: length/height-for-age, weight-for-age, weight-for-length, weight-for-height and body mass index-for-age: Methods and development. Geneva World Health Organ. 312 pages (2006).

26. Organization, W. H. & Fund (UNICEF), U. N. C. Recommendations for data collection, analysis and reporting on anthropometric indicators in children under 5 years old. (World Health Organization, 2019).

27. de Onis, M. et al. Comparison of the World Health Organization (WHO) Child Growth Standards and the National Center for Health Statistics/WHO international growth reference: implications for child health programmes. Public Health Nutr. 9, 942–947 (2006).

28. Garenne, M., Myatt, M., Khara, T., Dolan, C. & Briend, A. Concurrent wasting and stunting among under-five children in Niakhar, Senegal. Matern. Child. Nutr. 15, (2019).

29. Payandeh, A., Saki, A., Safarian, M., Tabesh, H. & Siadat, Z. Prevalence of Malnutrition among Preschool Children in Northeast of Iran, A Result of a Population Based Study. Glob. J. Health Sci. 5, 208–212 (2013).

30. Blencowe, H. et al. National, regional, and worldwide estimates of low birthweight in 2015, with trends from 2000: a systematic analysis. Lancet Glob. Health 0, (2019).

31. Ashorn, P. et al. Co-causation of reduced newborn size by maternal undernutrition, infections, and inflammation. Matern. Child. Nutr. 14, e12585 (2018).

32. Mertens, A., et al. (submitted). Causes and consequences of child growth faltering in low- and middle-income countries. (2020).

33. Perumal, N. et al. Effect of correcting for gestational age at birth on population prevalence of early childhood undernutrition. Emerg. Themes Epidemiol. 15, 3 (2018).

34. Barnett, A. G., van der Pols, J. C. & Dobson, A. J. Regression to the mean: what it is and how to deal with it. Int. J. Epidemiol. 34, 215–220 (2005).

35. Pardi, G., Marconi, A. M. & Cetin, I. Placental-fetal interrelationship in IUGR fetuses--a review. Placenta 23 Suppl A, S136-141 (2002).

36. Chou, F.-S. et al. Exposure to placental insufficiency alters postnatal growth trajectory in extremely low birth weight infants. J. Dev. Orig. Health Dis. 11, 384–391 (2020).

37. Levy, K., Woster, A. P., Goldstein, R. S. & Carlton, E. J. Untangling the Impacts of Climate Change on Waterborne Diseases: a Systematic Review of Relationships between Diarrheal Diseases and Temperature, Rainfall, Flooding, and Drought. Environ. Sci. Technol. 50, 4905–4922 (2016).

38. Saville, N. M. et al. Comprehensive analysis of the association of seasonal variability with maternal and neonatal nutrition in lowland Nepal. Public Health Nutr. 1–16 (2021) doi:10.1017/S1368980021003633.

39. Christian, P. et al. Risk of childhood undernutrition related to small-for-gestational age and preterm birth in low- and middle-income countries. Int J Epidemiol 42, 1340–1355 (2013).

40. Richard, S. A. et al. Wasting Is Associated with Stunting in Early Childhood123. J. Nutr. 142, 1291– 1296 (2012).

41. Khara, T., Mwangome, M., Ngari, M. & Dolan, C. Children concurrently wasted and stunted: A meta-analysis of prevalence data of children 6–59 months from 84 countries. Matern. Child. Nutr. 14, e12516 (2018).

42. Victora, C. G. et al. Revisiting maternal and child undernutrition in low-income and middle-income countries: variable progress towards an unfinished agenda. The Lancet 397, 1388–1399 (2021).

43. Myatt, M. et al. Children who are both wasted and stunted are also underweight and have a high risk of death: a descriptive epidemiology of multiple anthropometric deficits using data from 51 countries. Arch. Public Health 76, (2018).

44. Akombi, B. J., et al. Stunting, Wasting and Underweight in Sub-Saharan Africa: A Systematic Review. Int. J. Environ. Res. Public. Health 14, (2017).

45. Harding, K. L., Aguayo, V. M. & Webb, P. Factors associated with wasting among children under five years old in South Asia: Implications for action. PloS One 13, e0198749 (2018).

46. Mutunga, M., Frison, S., Rava, M. & Bahwere, P. The Forgotten Agenda of Wasting in Southeast Asia: Burden, Determinants and Overlap with Stunting: A Review of Nationally Representative Cross-Sectional Demographic and Health Surveys in Six Countries. Nutrients 12, 559 (2020).

47. Mutunga, M. et al. The relationship between wasting and stunting in Cambodian children: Secondary analysis of longitudinal data of children below 24 months of age followed up until the age of 59 months. PLOS ONE 16, e0259765 (2021).

48. Kohlmann, K. et al. Exploring the relationships between wasting and stunting among a cohort of children under two years of age in Niger. BMC Public Health 21, 1713 (2021).

49. Iversen, P. O., Ngari, M., Westerberg, A. C., Muhoozi, G. & Atukunda, P. Child stunting concurrent with wasting or being overweight: A 6-y follow up of a randomized maternal education trial in Uganda. Nutr. Burbank Los Angel. Cty. Calif 89, 111281 (2021).

50. Odei Obeng-Amoako, G. A., et al. Factors associated with concurrent wasting and stunting among children 6–59 months in Karamoja, Uganda. Matern. Child. Nutr. 17, e13074 (2020).

51. Moore, S. E. et al. Season of birth predicts mortality in rural Gambia. Nature 388, 434–434 (1997).

52. Rehkopf, D. H., Glymour, M. M. & Osypuk, T. L. The Consistency Assumption for Causal Inference in Social Epidemiology: When a Rose is Not a Rose. Curr. Epidemiol. Rep. 3, 63–71 (2016).

53. Prentice, A. M. & Cole, T. J. Seasonal changes in growth and energy status in the Third World. Proc. Nutr. Soc. 53, 509–519 (1994).

54. Rowland, M. G., Barrell, R. A. & Whitehead, R. G. Bacterial contamination in traditional Gambian weaning foods. Lancet Lond. Engl. 1, 136–138 (1978).

55. United Nations. Goal 2. Sustainable Development Knowledge Platform. https://sustainabledevelopment.un.org/sdg2.

56. Global action plan on child wasting: a framework for action to accelerate progress in preventing and managing child wasting and the achievement of the Sustainable Development Goals. https://www.who.int/publications/m/item/global-action-plan-on-child-wasting-a-framework-for-action.

57. ter Kuile, F. O., van Eijk, A. M. & Filler, S. J. Effect of sulfadoxine-pyrimethamine resistance on the efficacy of intermittent preventive therapy for malaria control during pregnancy: a systematic review. JAMA 297, 2603–2616 (2007).

58. Kayentao, K. et al. Intermittent preventive therapy for malaria during pregnancy using 2 vs 3 or more doses of sulfadoxine-pyrimethamine and risk of low birth weight in Africa: systematic review and meta-analysis. JAMA 309, 594–604 (2013).

## Supplementary References

1. McGrath, S., Zhao, X., Qin, Z. Z., Steele, R. & Benedetti, A. One-sample aggregate data meta-analysis of medians. Stat. Med. 38, 969–984 (2019).

2. Iqbal, N. T. et al. Promising Biomarkers of Environmental Enteric Dysfunction: A Prospective Cohort study in Pakistani Children. Sci. Rep. 8, 2966 (2018).

3. Ali, A. et al. Respiratory viruses associated with severe pneumonia in children under 2 years old in a rural community in Pakistan. J. Med. Virol. 88, 1882–1890 (2016).

4. Shrestha, P. S. et al. Bhaktapur, Nepal: The MAL-ED Birth Cohort Study in Nepal. Clin. Infect. Dis. 59, S300–S303 (2014).

5. Gladstone, B. P. et al. Protective Effect of Natural Rotavirus Infection in an Indian Birth Cohort. N. Engl. J. Med. 365, 337–346 (2011).

6. John, S. M. et al. Establishment of the MAL-ED Birth Cohort Study Site in Vellore, Southern India. Clin. Infect. Dis. 59, S295–S299 (2014).

7. Kattula, D., et al. The first 1000 days of life: prenatal and postnatal risk factors for morbidity and growth in a birth cohort in southern India. BMJ Open 4, e005404 (2014).

8. Ahmed, T. et al. The MAL-ED Cohort Study in Mirpur, Bangladesh. Clin. Infect. Dis. 59, S280–S286 (2014).

9. Kirkpatrick, B. D. et al. The “Performance of Rotavirus and Oral Polio Vaccines in Developing Countries” (PROVIDE) Study: Description of Methods of an Interventional Study Designed to Explore Complex Biologic Problems. Am. J. Trop. Med. Hyg. 92, 744–751 (2015).

10. Mduma, E. R., et al. The Etiology, Risk Factors, and Interactions of Enteric Infections and Malnutrition and the Consequences for Child Health and Development Study (MAL-ED): Description of the Tanzanian Site. Clin. Infect. Dis. 59, S325–S330 (2014).

11. Locks, L. M. et al. Effect of zinc and multivitamin supplementation on the growth of Tanzanian children aged 6–84 wk: a randomized, placebo-controlled, double-blind trial12. Am. J. Clin. Nutr. 103, 910–918 (2016).

12. Bessong, P. O., Nyathi, E., Mahopo, T. C. & Netshandama, V. Development of the Dzimauli Community in Vhembe District, Limpopo Province of South Africa, for the MAL-ED Cohort Study. Clin. Infect. Dis. 59, S317–S324 (2014).

13. Schoenbuchner, S. M. et al. The relationship between wasting and stunting: a retrospective cohort analysis of longitudinal data in Gambian children from 1976 to 2016. Am. J. Clin. Nutr. doi:10.1093/ajcn/nqy326.

14. Yori, P. P. et al. Santa Clara de Nanay: The MAL-ED Cohort in Peru. Clin. Infect. Dis. 59, S310–S316 (2014).

15. Jaganath, D. et al. First Detected Helicobacter pylori Infection in Infancy Modifies the Association Between Diarrheal Disease and Childhood Growth in Peru. Helicobacter 19, 272–279 (2014).

16. Bégin, F., Santizo, M.-C., Peerson, J. M., Torún, B. & Brown, K. H. Effects of bovine serum concentrate, with or without supplemental micronutrients, on the growth, morbidity, and micronutrient status of young children in a low-income, peri-urban Guatemalan community. Eur. J. Clin. Nutr. 62, 39–50 (2008).

17. Lima, A. A. M., et al. Geography, Population, Demography, Socioeconomic, Anthropometry, and Environmental Status in the MAL-ED Cohort and Case-Control Study Sites in Fortaleza, Ceará, Brazil. Clin. Infect. Dis. 59, S287–S294 (2014).

## Methods References

1. Organization, W. H. & Fund (UNICEF), U. N. C. Recommendations for data collection, analysis and reporting on anthropometric indicators in children under 5 years old. (World Health Organization, 2019).

2. Randev, S. Malnutrition in Infants under 6 months: Is it Time to Change Recommendations? Indian J. Pediatr. 87, 684–685 (2020).

3. WHO Multicentre Growth Reference Study Group. WHO child growth standards: length/height-for-age, weight-for-age, weight-for-length, weight-for-height and body mass index-for-age: Methods and development. Geneva World Health Organ. 312 pages (2006).

4. Richard, S. A. et al. Wasting Is Associated with Stunting in Early Childhood123. J. Nutr. 142, 1291–1296 (2012).

5. Home | Human Development Reports. https://hdr.undp.org/.

6. World Development Indicators | DataBank. https://databank.worldbank.org/source/world-development-indicators.

7. Benjamin-Chung, J. et. al. (submitted). Early childhood linear growth faltering in low-and middle-income countries. (2020).

8. Viechtbauer, W. Conducting Meta-Analyses in R with the metafor Package. J. Stat. Softw. 36, 1–48 (2010).

9. McGrath, S., Zhao, X., Qin, Z. Z., Steele, R. & Benedetti, A. One-sample aggregate data meta-analysis of medians. Stat. Med. 38, 969–984 (2019).

10. Cleveland, W. S. & Loader, C. Smoothing by Local Regression: Principles and Methods. in Statistical Theory and Computational Aspects of Smoothing (eds. Härdle, W. & Schimek, M. G.) 10–49 (Physica-Verlag HD, 1996). doi:10.1007/978-3-642-48425-4_2.

11. Ruppert, D., Wand, M. P. & Carroll, R. J. Semiparametric Regression. (Cambridge University Press, 2003). doi:10.1017/CBO9780511755453.

12. Wood, S. N., Pya, N. & Säfken, B. Smoothing Parameter and Model Selection for General Smooth Models. J. Am. Stat. Assoc. 111, 1548–1563 (2016).

13. Abatzoglou, J. T., Dobrowski, S. Z., Parks, S. A. & Hegewisch, K. C. TerraClimate, a high-resolution global dataset of monthly climate and climatic water balance from 1958–2015. Sci. Data 5, 170191 (2018).

14. Walsh, R. P. D. & Lawler, D. M. Rainfall Seasonality: Description, Spatial Patterns and Change Through Time. Weather 36, 201–208 (1981).

15. Papageorghiou, A. T. et al. The INTERGROWTH-21st fetal growth standards: toward the global integration of pregnancy and pediatric care. Am. J. Obstet. Gynecol. 218, S630–S640 (2018).

16. Cameron, N., Preece, M. A. & Cole, T. J. Catch-up Growth or Regression to the Mean? Recovery from Stunting Revisited. Am. J. Hum. Biol. 17, 412–417 (2005).

17. Barnett, A. G., van der Pols, J. C. & Dobson, A. J. Regression to the mean: what it is and how to deal with it. Int. J. Epidemiol. 34, 215–220 (2005).

18. Linden, A. Assessing regression to the mean effects in health care initiatives. BMC Med. Res. Methodol. 13, 119 (2013).

